# Polygenic effect on accelerated tau pathology accumulation in Alzheimer’s disease: implications for patient selection in clinical trials

**DOI:** 10.1101/2021.11.02.21265788

**Authors:** Anna Rubinski, Simon Frerich, Rainer Malik, Nicolai Franzmeier, Alfredo Ramirez, Martin Dichgans, Michael Ewers, Alzheimer’s Disease Neuroimaging Initiative (ADNI)

## Abstract

Progression of fibrillar tau is a key driver of dementia symptoms in Alzheimer’s disease (AD), but predictors of the rate of tau accumulation at patient-level are missing. Here we combined the to-date largest number of genetic risk variants of AD (n=85 lead SNPs) from recent GWAS to generate a polygenic score (PGS) predicting the rate of change in fibrillar tau. We found that a higher PGS was associated with higher rates of PET-assessed fibrillar-tau accumulation over a mean of 1.8 yrs (range = 0.6 – 4 yrs). This, in turn, mediated effects of the PGS on faster rates of cognitive decline. Sensitivity analysis showed that the effects were similar for men and women but pronounced in individuals with elevated levels of beta-amyloid and strongest for lead SNPs expressed in microglia. Together, our results demonstrate that the PGS predicts tau progression in Alzheimer’s disease, which could afford sample size savings by up to 34% when used alone and up to 61% when combined with APOE ε4 genotype in clinical trials targeting tau pathology.

## Introduction

Alzheimer’s disease (AD) is the major cause of dementia with an estimated 50 million cases worldwide ^1^. The development of tau pathology is a disease-defining pathology that drives cognitive decline in AD ^2^. The rates of tau-accumulation and associated symptomatic worsening vary substantially between patients ^3,4^; however, little is known about those factors that determine the rate of tau progression in AD ^5-7^, thus hampering the assessment of treatment efficacy in ongoing clinical trials and the prognosis of dementia in clinical praxis.

Here, we propose to employ a polygenic score (PGS) for the prediction of tau progression in AD, which could be utilized to select individuals with faster tau progression in clinical trials. The PGS is a powerful tool to assess an individual’s genetic risk for AD by integrating the effects of multiple single-nucleotide polymorphisms (SNPs) discovered in GWAS ^8^. Previous studies demonstrated that PGSs show utility for the prediction of AD risk ^9-11^ and age at dementia onset ^11-13^. However, mixed evidence has been reported for PGS as a predictor of the rate of cognitive worsening ^11,14^. It also remains unclear whether a PGS is predictive of the progression of tau pathology, which underlies cognitive decline. So far, studies examining the value of a PGS for predicting tau pathology have been limited to cross-sectional assessments ^13,15-17^, leaving the question unaddressed whether a PGS is associated with faster rates of fibrillar tau progression ^18^.

Against this background, our primary aim was to investigate the value of PGS for the prediction of longitudinal changes in fibrillar tau and thus the rate of cognitive worsening. Our secondary aim was to test the generalizability of the abovementioned association between PGS and the progression of fibrillar tau. In light of previous studies showing a strong association between elevated beta-amyloid (Aβ) levels and higher cortical fibrillar tau ^19,20^ we primarily focused on Aβ as a modulating factor. Because previous PGSs were associated with higher Aβ ^15,17,21-25^, we further tested whether the present PGS is associated with faster fibrillar tau accumulation merely as a consequence of the effect on Aβ, or by interacting with Aβ and thus modulating the effects of Aβ on fibrillar tau development. This is of interest for risk stratification strategies in clinical trials, because an interaction between PGS and Aβ for predicting tau progression would render the PGS an important factor that could inform Aβ-based risk enrichment in clinical trials on AD. Given previous reports of sex-dependent effects of genetic AD-risk variants on tau-pathology ^26,27^, we further assessed whether sex modulated the effects of the PGS on tau-PET. Finally, we investigated whether the predictive value of the PGS on tau progression and cognitive decline is cell-type dependent. This was motivated by previous studies suggesting that AD risk genes are preferentially expressed in microglia ^28,29^. We therefore computed cell-type-specific PGSs by combining lead SNPs whose genes are expressed in the same cell type. We thus generated PGS for seven major cell types, and tested whether the PGSs are predictors of faster progression in tau-PET accumulation.

For the present work, we leveraged two recently published GWAS that included up to 1.1 million participants ^30,31^ and more than doubled the number of known independent AD risk variants. Hence, we used 85 lead variants to calculate a novel PGS. We compared the predictive power of this novel PGS to that of the APOE ε4 genotype, which is the strongest genetic risk factor of AD dementia ^32^. We tested the genetic associations in deeply phenotyped individuals who were longitudinally assessed with neuropsychological testing and molecular PET tracers of fibrillar tau and Aβ in the Alzheimer’s Disease Neuroimaging Initiative (ADNI) ^33^, one of the world’s largest multicenter biomarker studies on AD. Overall, our findings answer the question whether the PGS is not only associated with higher AD risk but also with higher tau progression during the course of AD. Our results demonstrate utility of the PGS for risk enrichment and thus a reduced sample size in clinical trials.

## Results

### The PGS is associated with faster accumulation rates of tau and cognitive decline

We computed a PGS based on 85 independent lead SNPs from two recent GWAS ^30,31^, excluding APOE ε4 genotype (for lead SNPs see **Supplementary Table 1**, for PGS distribution see **Supplementary Figure 1)**. Using linear mixed effects models, we estimated the individual rates of change in tau-PET obtained from three *a priori* established composite ROIs as defined by the Braak post-mortem staging of tau pathology (**Figure 1A**). We tested the PGS as a predictor of the estimated rates of change in tau-PET in each ROI, controlling for age, sex, education, diagnosis, ethnicity, APOE genotype, and 10 principal components of the genetic population structure (see Methods).

**Table 1:** Sample characteristics

|  | **Tau-PET**<br>(n=231) | **Aβ-PET**<br>(n=879) | **MEM**<br>(n=1832) | **ADAS13**<br>(n=1813) |
| --- | --- | --- | --- | --- |
| Age | 74.67 [58 - 92] | 73.03 [55 – 94] | 73.59 [55 – 91] | 73.61 [55 – 91] |
| Sex (M/F) | 119M / 112F | 450M / 429 F | 1002M / 830 F | 991M / 822F |
| Diagnosis<br>(HC/MCI/AD) | 135HC/ 74MCI/<br>22AD | 417HC/ 399MCI/<br>63AD | 646HC/ 870MCI/<br>316AD | 643HC/ 868MCI/<br>302AD |
| Aβ status<br>(-/+/na) | 103/ 126/ 2 | 489/ 390/ 0 | 646/ 802/ 384 | 641/ 790/ 382 |
| APOE ε4 (-/+) | 127/ 104 | 531/ 348 | 986/ 846 | 977/ 836 |
| Ethnicity<br>(% White) | 89.6% | 89.8% | 90.2% | 90.2% |
| Follow-up<br>time, y | 1.78 [0.65 - 3.95] | 4.17 [0.86 - 9.33] | 4.09 [0.36 - 15.03] | 4.2 [0.34 - 15.12] |

Aβ = beta-amyloid; ADAS = Alzheimer's Disease Assessment Scale; APOE = apolipoprotein E; MEM = episodic memory, N = sample size.
Unless otherwise indicated, the summary statistics are presented as mean [range]

**Figure 1:**
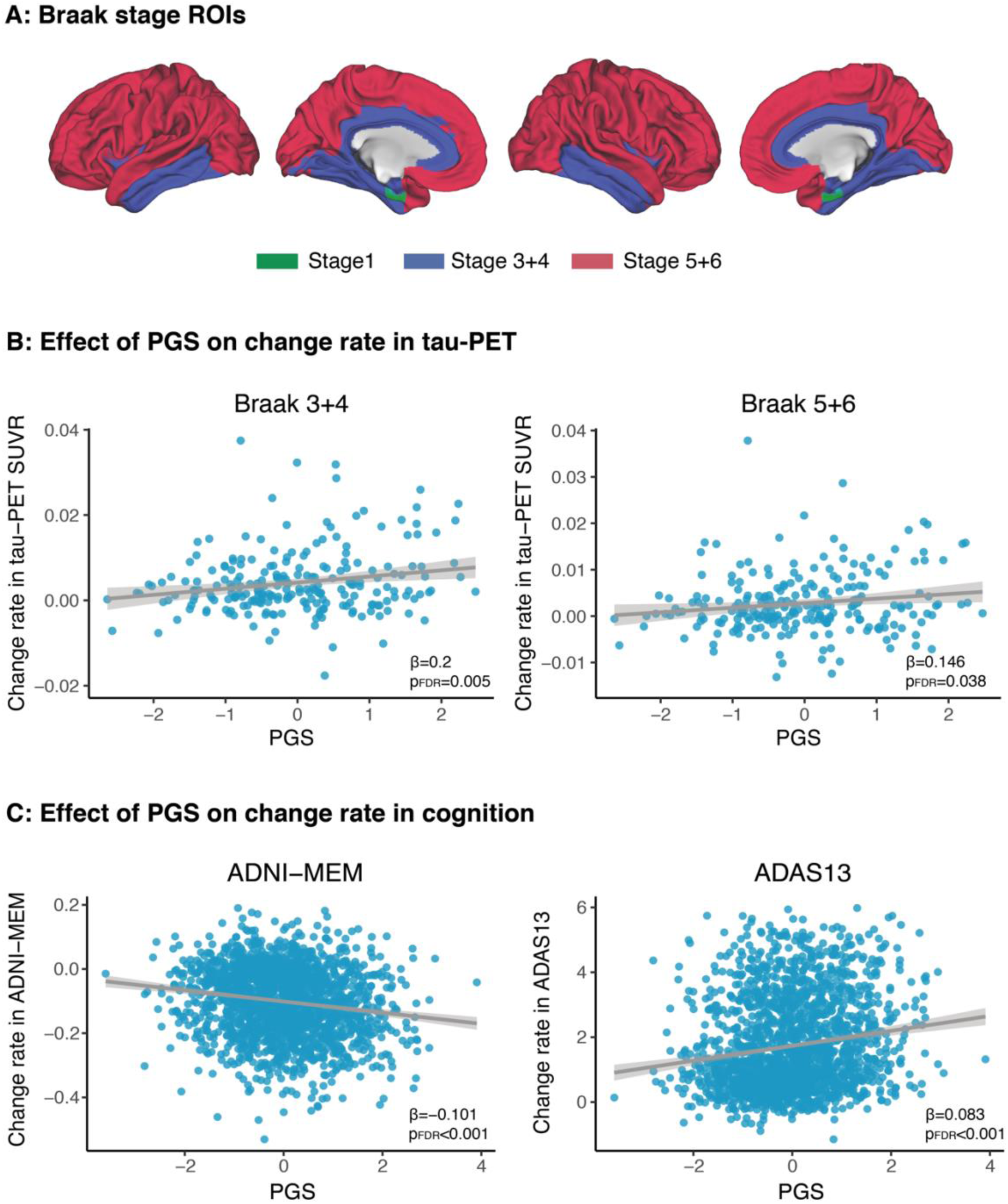
Spatial mapping of Braak stage-specific ROIs and the association between PGS and rate of change in tau-PET and cognition. Surface rendering of the composite Braak-stage ROIs ^93^ that were used to determine regional tau-PET uptake. Braak stage 2 (hippocampus) was not included due to the off-target binding of the AV1451 tau-PET tracer **(A)**. Scatterplots showing the regression line (solid blue) and 95% CI (shaded area) for the associations between PGS and the rate of change in tau-PET SUVRs **(B)** and cognition **(C)**. Standardized β-values and FDR corrected p-values are shown. ADAS13 = Alzheimer’s Disease Assessment Scale cognitive subscale; ADNI-MEM = Alzheimer’s Disease Neuroimaging Initiative memory composite; PGS = Polygenic score.

A higher PGS was associated with higher accumulation rates of tau-PET in cortical regions (Braak 3+4 ROI: β=0.2, pfdr=0.005; Braak 5+6 ROI: β=0.146, pfdr=0.038; **Figure 1B**), but not in entorhinal Braak 1 ROI (pfdr= 0.215). A higher PGS was further associated with a faster decline in episodic memory as assessed by the composite score ADNI-MEM (β=-0.101, pfdr<0.001; **Figure 1C**) and global cognitive performance as assessed by ADAS13, i.e. the extended cognitive subscale of the Alzheimer’s Disease Assessment Scale (ADAS) (β=0.083, pfdr<0.001; **Figure 1C**) over a period of 4 years on average (range = 0.4 – 15 years). Bootstrapped mediation analysis showed that higher rates of global tau-PET accumulation mediated the effect of the PGS on the rate of change in ADNI-MEM (mediation effect: β=- 0.004 [95% CI: -0.007, -0.001], p=0.010, proportion mediated = 30.7%; **Figure 2A**) and ADAS13 (mediation effect: β=0.041 [95% CI: 0.012, 0.08], p=0.006, proportion mediated = 23.2%; **Figure 2B**), suggesting that the effect of the PGS on the rate of tau-PET explains the association between a higher PGS and faster cognitive decline.

**Figure 2:**
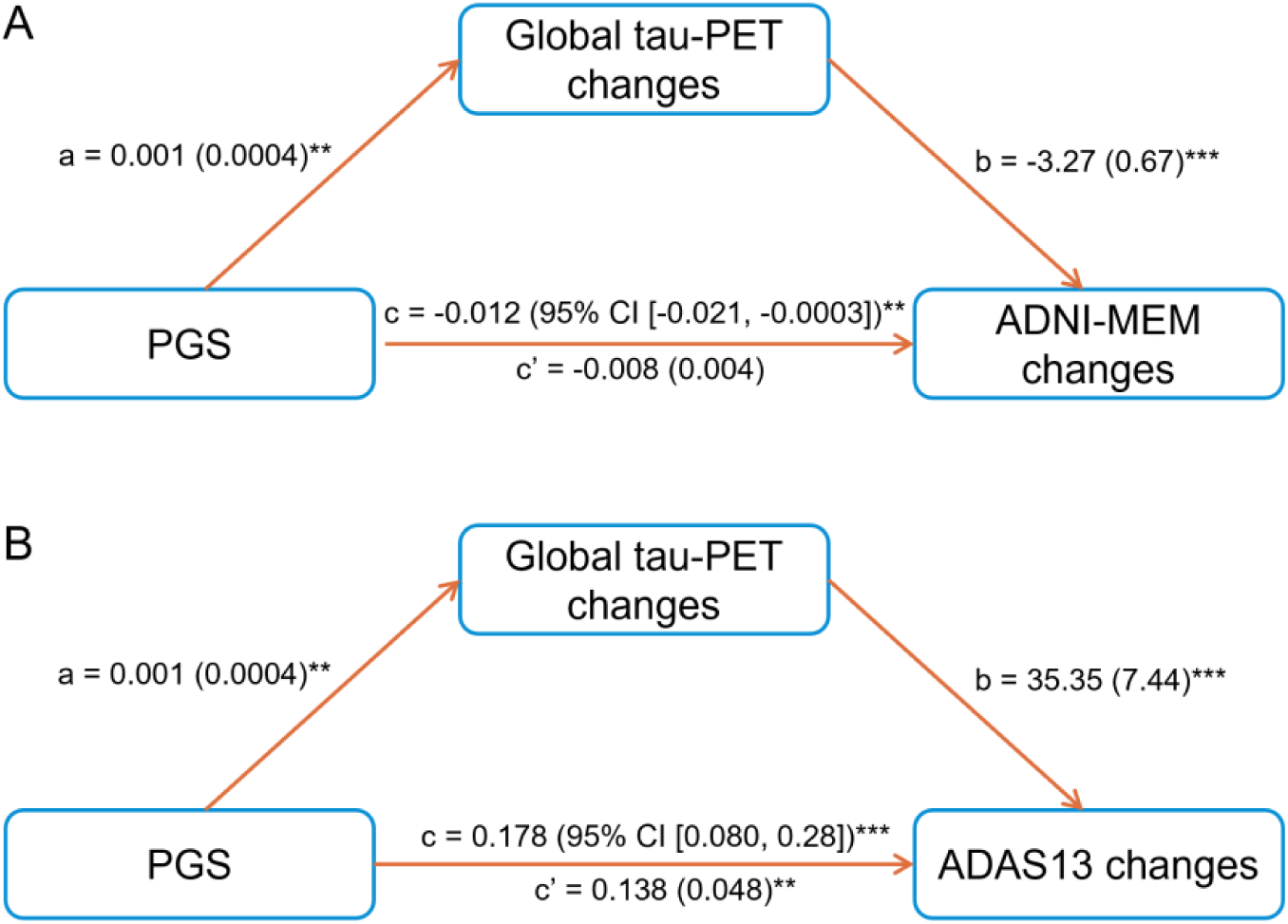
The rate of change in global tau-PET mediates the effect of PGS on change rate in cognition. Path model illustrating the mediation analyses. The effect of PGS on the rate of change in global tau-PET changed mediated the effect of PGS on ADNI-MEM **(A)** and ADAS13 **(B)**. Path weights are displayed as β-values with standard errors displayed in brackets. The path weight *c* indicates the effect of PGS on cognitive measures (i.e. either ADNI-MEM or ADAS13) without taking global tau-PET changes into account, the path coefficient *c’* indicates the corresponding effects of PGS after accounting for the mediator including global tau-PET changes. The total effect was determined using bootstrapping with 1000 iterations. Models were controlled for age, sex, education, APOE genotype, diagnosis, PC1-10, and ethnicity. ADAS13 = Alzheimer’s Disease Assessment Scale cognitive subscale; ADNI-MEM = Alzheimer’s Disease Neuroimaging Initiative memory composite; PGS = Polygenic score.

For the sake of completeness, we also tested the association between the PGS and cross-sectional levels of tau-PET and cognition in an analogous way and found a higher PGS to be associated with higher tau-PET levels and lower memory as well as lower global cognitive scores (**Supplementary Figure 2; see Supplementary Table 3 for statistics**).

### Aβ levels do not mediate but modulate the PGS effect on tau-PET increases

First, we replicated previous findings of associations with increased Aβ ^25^, observing that a higher PGS was associated with faster rates of global amyloid-PET accumulation (β=0.069, p^FDR^=0.038). Next, we tested whether the effect of the PGS on tau-PET is independent of changes in amyloid-PET. When controlling for the rate of change in amyloid-PET in addition to the other covariates, the effects of the PGS on the rate of change in tau-PET remained significant in Braak stage 3+4 (β=0.149, p=0.04), suggesting that the association between the PGS and increases in amyloid-PET does not account for the association between the PGS and tau-PET changes.

However, we found evidence for more pronounced effects of the PGS in presence of elevated levels of Aβ. When subjects were stratified into those with abnormal levels of amyloid-PET (Aβ+) versus those with normal levels of amyloid-PET (Aβ-), we found that the effects of the PGS on the rate of tau-PET accumulation in cortical areas (Braak-stage 3-6 regions) were significant in the Aβ+ but not Aβ- participants (**Figure 3A; See supplementary Table 4 for sample characteristics and supplementary Table 5 for statistics**), indicating that the PGS is associated with enhanced fibrillar tau accumulation in the presence of AD-like abnormal Aβ levels but not in normal aging.

**Figure 3:**
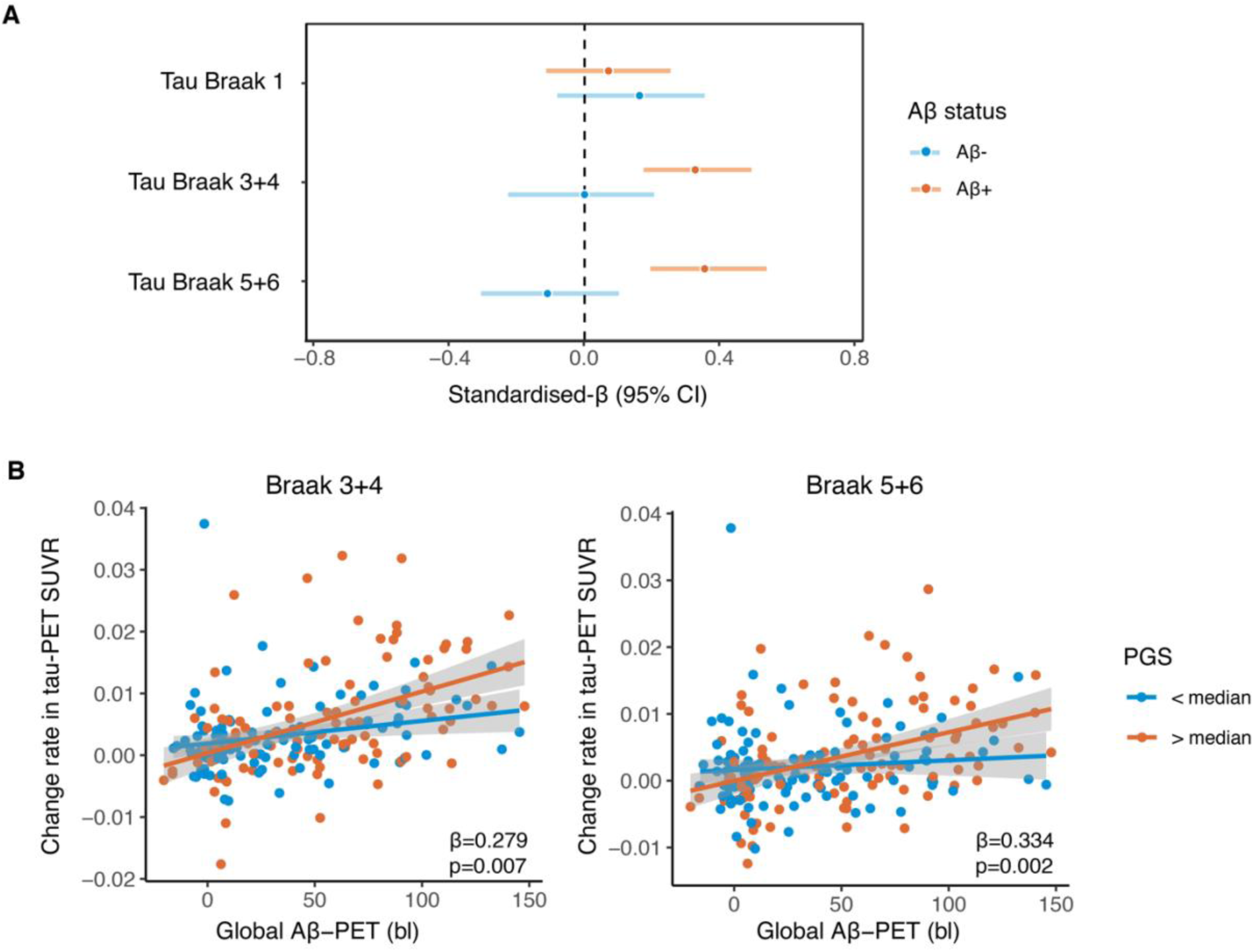
PGS modulates the effect of amyloid on change rate in tau-PET. Regression weights of the association between the PGS and change rate in tau-PET SUVRs stratified by amyloid-PET status (Aβ+ (red color) vs. Aβ- (blue color)) are shown **(A)**. Weights are indicated as standardized β-values with 95% confidence intervals (CI) derived from 1,000 bootstrapping iterations. Scatterplots showing the rate of change in tau-PET SUVRs as a function of amyloid-PET centiloid **(B)**. Regression lines are shown for the group including individuals with PGS values > median (red) and for the group with PGS < median (blue); the shaded area represents the 95%CI. Note that PGS levels were stratified to high and low (median split) only for illustrational purposes, whereas the regression analyses to estimate the regression weights was computed based on the predictor PGS as a continuous variable.

Given that in Aβ+ subjects, higher abnormal levels of Aβ are a strong driver of fibrillar tau accumulation in the cortex ^3^, we tested whether the PGS modulates Aβ-related increases in the rate of fibrillar tau accumulation. In regression analysis, we found a significant interaction of the PGS by global amyloid-PET accumulation on subsequent increase in tau-PET (Braak 3+4: interaction term β=0.279, p=0.007; Braak 5+6; interaction term β=0.334, p=0.002; **Figure 3B**). Higher levels of the PGS were associated with a stronger effect of amyloid-PET on subsequent rates of tau-PET increases. Together, these results suggest that the PGS shows a synergistic effect with elevated levels of Aβ on the rate of tau-PET increases such that the PGS is associated with enhanced tau-PET accumulation particularly in individuals with higher abnormal levels of Aβ.

### No sex-dependent effects of the PGS on tau-accumulation

We next investigated whether sex modifies the association between the PGS and the rate of tau-PET changes. Neither in the whole sample nor in an analysis stratified by Aβ status, the interaction effect of the PGS by sex on the rate of tau PET accumulation was significant (p > 0.05).

### Cell-type-specific PGS associations with tau-accumulation

To test whether the association between the PGS and tau-PET accumulation varies by cell type, we computed cell-type-specific PGSs. These scores only contained AD risk variants whose genes are expressed in the respective cell type, including neuronal cells (GABAergic, glutamatergic), microglia, oligodendrocytes, oligodendrocyte precursor cells (OPC), astrocytes, endothelial cells (see **Supplementary Table 2** for the SNPs included in each cell-type-specific PGS).

**Table 2:** Estimated sample size required for detecting intervention effects on tau-PET changes in the full sample and in a subgroup of Aβ+ individuals at power = 0.8

| Group | Subgroup | Required number of participants per arm<br>to detect intervention effect of |  |
| --- | --- | --- | --- |
|  |  | 20% | 40% |
| All | All participants | 630 | 144 |
| | APOE ε4+ | 446 (29%) | 96 (33%) |
|  | 4th PGS quartile | 425 (33%) | 95 (34%) |
| | 4th PGS quartile + APOE ε4+ | 231 (63%) | 50 (65%) |
| Aβ + | All Aβ + participants | 440 | 97 |
| | APOE ε4+ | 374 (15%) | 80 (18%) |
|  | 4th PGS quartile | 290 (34%) | 66 (32%) |
| | 4th PGS quartile + APOE ε4+ | 176 (60%) | 38 (61%) |

Percentage in brackets indicates the size of the sample size reduction relative to the reference group of no stratification (all participants) within either whole group (All) or the Aβ + group. APOE ε4+ = APOE $\epsilon$ 3/ ε4 & APOE ε4/ ε4 carrier, 4<sup>th</sup> PGS quartile = group in upper quartile of residualized PGS scores, Aβ + = abnormally high amyloid-PET accumulation

We found that exclusively the microglial PGS was associated with faster tau-PET accumulation in Braak-stage 3+4 ROI (p=0.04, adjusted for multiple comparisons; **Figure 4, Supplementary Table 6**). Nominal associations between a higher oligodendrocyte PGS and faster tau-PET accumulation in Braak-stage 3+4 and 5+6 ROIs were observed, which however did not survive correction for multiple comparisons (**Figure 4, Supplementary Table 6**). For cognitive performance, we found a higher PGS of each cell type to be associated with faster worsening in episodic memory and global cognition (**Figure 4, Supplementary Table 6**), suggesting cell-type independent effects on the cognitive endophenotype. For amyloid-PET changes, an association between a higher microglial PGS and higher rate of amyloid-PET was observed at the nominal significance level, which did not survive correction for multiple comparisons (**Figure 4, Supplementary Table 6**).

**Figure 4:**
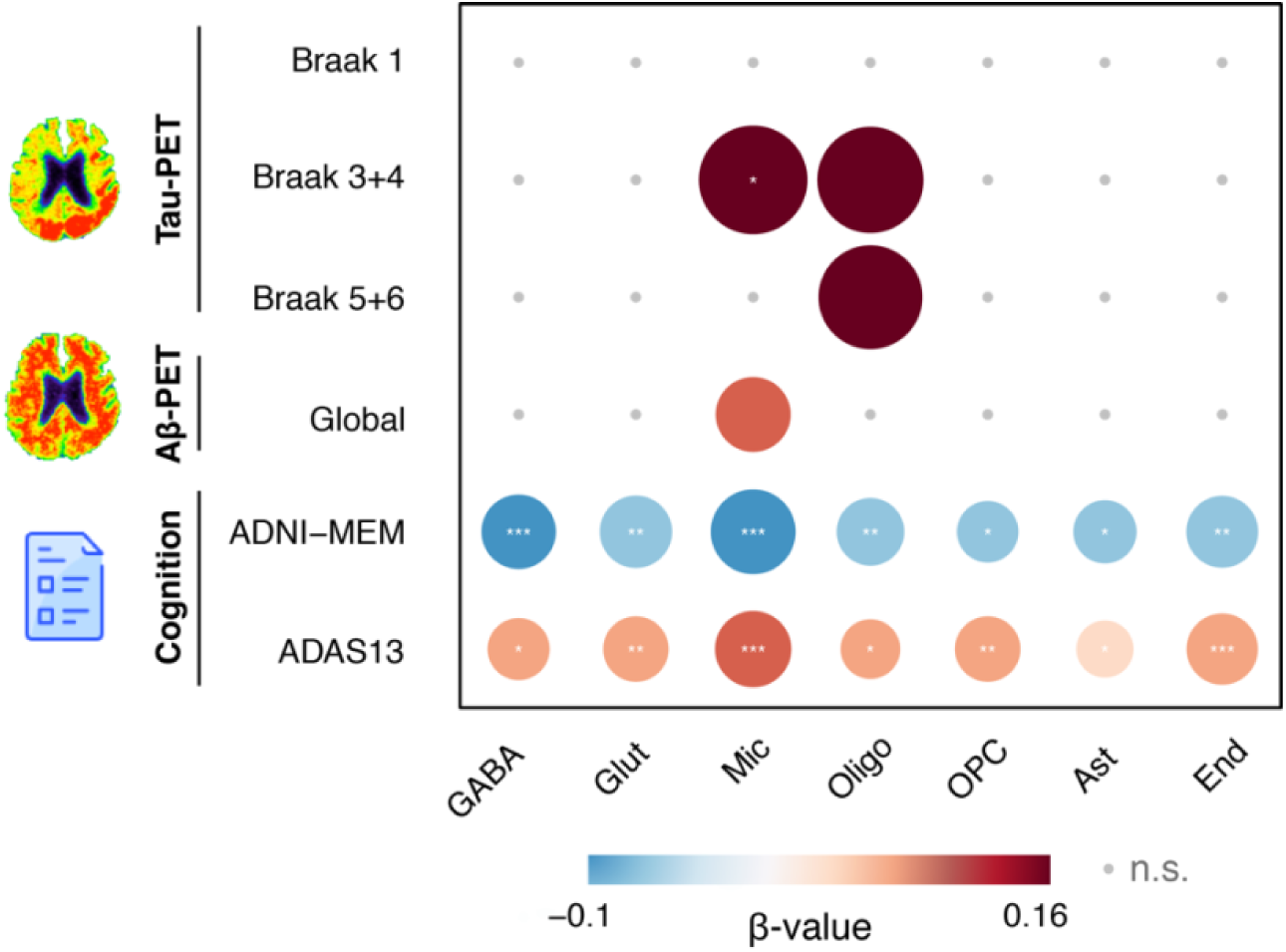
Association between cell-type-specific PGS and the rate of changes in tau-PET, amyloid-PET and cognition. Tabulation of the effects of cell-type-specific PGS on each outcome measure. The magnitude of the standardized β-values is represented by the circle size and the direction of the association is represented by color (red for positive associations and blue for negative associations). Significant associations after FDR correction are represented by an asterisk (*pFDR<0.05; **pFDR<0.01; ***pFDR<0.001). Cell-type-specific scores correspond to the following cell types: GABA = GABAergic neurons; Glut = glutamatergic neurons; Mic = microglia; Oligo = oligodendrocytes; OPC = oligodendrocyte precursor cells; Ast = astrocytes; End = endothelial cells.

### PGS increases sensitivity to detect tau-targeting intervention effects

Finally, we tested the utility of a higher PGS for risk enrichment in clinical trials targeting fibrillar tau. To this end, we estimated the sample size needed to detect hypothetical treatment effects that reduced the rate of increase in tau-PET by 20% and 40% when stratified by PGS scores (lowest versus highest quartile of PGS). We estimated the PGS effects on the sample size needed for the whole group and in the group restricted to Aβ+ individuals. For a clinical trial on tau-PET changes regardless of Aβ status, results showed that the PGS stratification yielded a saving in sample size by 33 - 34 % compared with the no-stratification scenario, depending on the size of the assumed treatment effect (**Table 2**). For a clinical trial restricted to Aβ+ subjects, stratification by PGS yielded a reduction in sample size needed by 32 – 34% compared to no stratification in the Aβ+ subjects. Previous studies found the APOE ε4 genotype to be associated with faster rates of tau-PET increases in AD ^5^. When compared to the stratification based on APOE ε4 status (presence of absence of APOE ε4 allele), there was a substantial advantage of PGS stratification, where the reduction in sample size based on APOE ε4 status was only half of that attained when based on the PGS for stratification in the Aβ+ group. Furthermore, both APOE status and a higher PGS showed synergistic effects, where risk enrichment based on both variables reached between 60 - 61% of reduction in sample size in the Aβ+ group (**Table 2**).

Given our findings on cell-type-specific PGSs, we further estimated the utility of the microglia PGS for risk stratification. Results showed that the microglia PGS yielded – compared to the total PGS – a comparable saving in sample size in the Aβ+ group, but a numerically lower sample-size reduction in the whole group (**Supplementary table 7**).

## Discussion

Here, we combined longitudinal molecular PET with a comprehensive set of lead SNPs from the to-date largest GWAS on AD for the polygenic prediction of the rate of fibrillar tau progression and cognitive changes. Our primary finding showed that a higher PGS was associated with faster rates of tau-PET over an average of 2 years of follow-up assessment in Aβ+ individuals, which mediated the PGS effect on faster cognitive decline. These results demonstrate that the accelerated increase in pathologic tau accumulation underlies the association between the PGS and symptomatic worsening in AD. Our findings support the utility of the PGS for risk enrichment in clinical trials on tau-PET changes, which may yield substantial savings in sample size needed to test treatment effects on the progression of tau pathology in AD.

Our study makes important contributions towards a clinically relevant PGS-based prediction of increases in tau pathology and thus cognitive decline in AD. We demonstrated for the first time that a PGS predicts longitudinal changes in fibrillar tau as assessed by tau-PET, i.e. the best-established biomarker of fibrillar tau pathology recently adopted as outcome measure in several clinical trials on AD ^34^. Our results suggest that the value of the PGS for the prediction of tau-PET changes translates into a reduction of the required sample size by up to 34% in clinical trials targeting tau pathology. Importantly, when applied in Aβ+ groups, risk stratification by the PGS more than doubles the sample-size reduction compared to that by the APOE-ε4 based stratification, supporting the added value of the PGS for the prediction of tau progression in clinical trials. The development of powerful predictors for risk stratification is a pressing need, since the rate of change per year in tau-PET ranges between 0.5 and 3% in cognitively normal Aβ+ subjects and 3 – 8% in symptomatic individuals ^35^. Thus, in a clinical trial targeting tau, the control group would show a substantial variability of tau-PET changes over 2 years of follow-up, rendering feasible intervention effects on tau difficult to detect. Therefore, the PGS could be of great value to identify subjects of imminent worsening of tau accumulation in clinical trials and may support individualized disease management within a precision medicine guided strategy ^8^.

Although a PGS can be constructed based on a larger number of SNPs selected at a more liberal p-value ^36^, we focused our analysis on genome-wide significant lead SNPs. Compared to a PGS that relies on a broader genetic background, this approach has key advantages regarding clinical applications. First, it facilitates establishing cut-off values for risk stratification. Second, it enhances comparability between studies and the interpretability of the PGS that could be compromised when including a larger number of SNPs with potentially spurious associations

^36,37^. A strength of our study is the addition of over 42 novel lead SNPs beyond those already known for computing the PGS ^30,31^, thus enhancing the statistical power for testing the PGS as a predictor of tau-PET and cognitive changes. We note that although the effect of a PGS derived from AD-risk variants on prediction of longitudinal cognitive worsening has been previously supported ^25,30,38-40^, it has not been exempt of conflicting evidence ^41-43^. This discrepancy of results might derive from a limited sample size and lower number of risk variants included in previous PGSs ^41-43^. Also, it is unclear to what extent those PGSs were associated with faster changes in fibrillar tau progression. We demonstrate that the predictive power of the present PGS is dependent on the effect of tau-PET changes. Overall, the current study substantially adds to previous studies that used a lower number of genome-wide significant SNPs for construing a PGS and were confined to cross-sectional analyses of pathologic tau ^15,16,^for review see: ^18,44^.

Our secondary aim was to examine possible factors that moderate the association between the PGS and tau progression, primarily focusing on Aβ deposition. Longitudinal studies showed that the presence of elevated levels of Aβ in the cortex is a major driver of cortical increases in fibrillar tau in AD ^3,19^. Therefore, one possibility is that the effect of the PGS on tau-PET changes is merely indirectly caused through a PGS-related increase in Aβ accumulation. However, we demonstrated that the PGS effect on tau-PET changes in cortical brain regions *cannot* be reduced to an indirect effect of PGS on levels of amyloid-PET: controlling for amyloid-PET did not diminish the PGS effects on tau-PET in Braak stage 3+4. Rather, the effects of the PGS on the rate of tau-PET accumulation were pronounced in Aβ+ individuals. Furthermore, in the Aβ+ individuals we observed a cross-talk between PGS and baseline levels of amyloid-PET on the rate of subsequent change in tau-PET, suggesting that the PGS affects the formation of tau pathology in cortical brain regions downstream of the abnormally elevated Aβ. In contrast, previous studies on APOE ε4 status reported that the effect of APOE ε4 on higher cortical tau-PET accumulation were due to the effect on increased levels of Aβ ^45-47^. Taken together, these results suggest different roles of the APOE ε4 allele and PGS for the prediction of tau-PET changes, where presence of the APOE ε4 allele contributes to cortical tau-progression indirectly via increasing levels of Aβ, but it is the PGS that is associated with faster rates of tau-accumulation once abnormal levels of Aβ are present.

We did not find sex to modulate the effect of the PGS on the rate of tau-PET changes, despite previous reports of sex differences in the risk of AD ^48^. We caution that our results do not implicate the absence of any sex-dependent effects of genetic risk variants of AD on tau pathology, rather the cumulative polygenic effects on the progression of tau pathology appear to be similarly distributed across both sexes. The strongest evidence for sex-dependent genetic effects on tau-pathology exists for APOE ε4 genotype, where the association between APOE ε4 and tau-accumulation is stronger in females compared to males ^26,27,49^. The current study controlled for APOE ε4 genotype, and is therefore not in conflict with these previous findings. Few studies employed sex-stratified GWAS or family-based association studies in AD, reporting sex-specific risk variants associated with AD risk ^50^ and higher levels of Aβ and tau pathology ^51^. Notably, a subset of sex-specific SNPs overlapped with previously established AD risk variants, some more pronounced in females, others in males ^51-53^. Due to the large number of SNPs included in the current PGS, we assume that potential sex-dependent biases towards either males or females at single-SNP level were balanced out. Still, future studies may establish sex-dependent PGSs for the prediction of pathologic tau progression ^53^.

Lastly, our cell-type-specific PGSs suggested a selective effect of the microglial PGS on the rate of tau-PET accumulation, whereas the oligodendrocyte PGS showed only a nominal association that did not survive multiple comparison correction. Our results are consistent with previous GWAS-related studies implicating a crucial role in particular of microglial alterations in the etiology of AD pathology ^17,29,54-56^. Previous gene set analyses showed immune response to be the predominant pathway linked to AD risk ^31,56,57^, and microglia are altered in association with tau pathology ^58^. Notably, when excluding SNPs within the APOE region, immune-related pathways showed the strongest associations ^56,59^. In line with a strong enrichment of AD-related genes in microglia ^31^, we show that a microglial cell-type-specific PGS is associated with faster tau-PET accumulation, supporting a key role of microglial alterations in the generation of tau pathology. Microglia-targeting treatments such as antibody-mediated activation of TREM2 are underway ^60^, where cell-type-specific PGS could make a unique contribution within a precision medicine guide strategy to identify those subjects with immune response alterations linked to AD risk ^59,61^.

For the interpretation of our findings, several limitations of our study should be considered. First, lead SNPs were obtained from GWAS that were conducted mostly in European-ancestry individuals ^30,31,62^. Furthermore, ADNI participants included in the current study are primarily self-reporting as white. We caution that the results may not generalize to individuals of other ethnic background, and recognize that need for more research individuals from underrepresented ethnic and racial background ^63^. Second, our PGS was limited to common SNPs with a minor allele frequency of >0.01, and potential epistatic and gene-environment interactions were not considered. Third, an inflation of our findings due to a partial sample overlap cannot be excluded, as the lead SNPs included in the current analysis were derived from previous meta-analytic GWAS on AD risk that partially included samples from ADNI ^30^. Lastly, we did not investigate any potential differences between lead SNPs in the contribution to the prediction of tau accumulation. However, both the predictive value and pathomechanistic pathways likely vary between different lead SNPs ^64^. Previous studies focusing on particular lead SNPs showed associations for genetic variants in genes including BIN1 ^65,66^, among others ^51,67^ to be associated with alterations in tau-pathology in AD. However, the small effect size attributed to each risk variant reduces its potential predictive value on a specific phenotype like contribution to tau pathology. Therefore, the current study focused on the cumulative effects across a larger set of SNPs for the prediction of the rate of tau pathology in order to maximize the predictive power.

In conclusion, the present PGS is a promising tool for the prediction of the rate of tau-progression that will enhance risk enrichment in clinical trials and potentially inform therapeutic decision for treating tau pathology in AD. Future studies will be needed in order to test potential interactions between the PGS and other known risk factors of AD such as life style factors and cerebrovascular changes ^68,69^. Furthermore, presence of protective SNPs in genes such as Klotho ^70^, PLCG2 ^71^ may somewhat compensate the effects of a PGS on tau-progression. Thus, the current study encourages future studies to explore additional predictors towards establishing a comprehensive and cost-effective prediction model for the progression of tau pathology.

## Methods

We followed the recently developed Polygenic Risk Score Reporting Standards ^72^ (PRS-RS, see **Supplementary table 8** for a checklist).

### ADNI participants

All data were obtained from the Alzheimer’s Disease Neuroimaging Initiative (ADNI) database (adni.loni.usc.edu). ADNI is a longitudinal multicenter study designed to develop clinical, imaging, genetic, and biochemical biomarkers for the early detection and tracking of Alzheimer’s disease ^33^. Participants were selected based on the availability of genotype data (accessed on 19/01/2021) and at least two measurements of tau-PET, amyloid-PET or cognition (for follow-up duration see **Table 1**). Ethnicity was determined based on self-report.

Clinical classification was performed by ADNI centers as cognitively normal (CN, Mini Mental State Exanimation (MMSE) ≥ 24, Clinical Dementia Rating (CDR) = 0, no memory concerns), amnestic mild cognitive impairment (MCI, MMSE ≥ 24, CDR = 0.5, subjective memory concern, objective memory loss measured by education adjusted scores on the Wechsler Memory Scale Memory II, absence of significant levels of impairment in other cognitive domains, essentially preserved activities of daily living and an absence of dementia) or AD dementia following standard diagnostic criteria ^73^.

Ethical approval was obtained by the ADNI investigators, all participants provided written informed consent (further information about the inclusion/exclusion criteria may be found at www.adni-info.org).

### Genotyping procedures and quality control in ADNI

The missing genotypes in ADNI were first imputed based on the HRC reference panel v1.1 ^74^ using Minimac4 on the Michigan imputation server ^75^, separately by cohort, as ADNI genotypes were genotyped on three different Illumina arrays, namely Human610-Quad (620,901 markers), HumanOmniExpress (730,525 markers), and HumanOmni2.5-8 (2,379,855 markers). Prior to the imputation, strand, positions, and ref/alt assignments were updated if inconsistent between ADNI genotypes and the HRC reference panel. SNPs were removed if they had differing alleles, inconsistent allele frequencies (>0.2 difference), no equivalent in the reference panel, or if they were palindromic SNPs with a minor allele frequency (MAF) >0.4 (via www.well.ox.ac.uk/~wrayner/tools/HRC-1000G-check-bim-v4.3.0.zip). SNPs with an imputation r^2^ <0.5 were excluded.

### Calculation of genetic scores

#### Genetic risk variants

We considered variants associated with AD or AD related dementias (ADD) at a genome-wide significant level of p≤5×10^−^8 in the most recent and largest AD/ADD GWAS including 111,326 cases/677,663 controls and 90,338 cases/1,036,225 controls, respectively ^30,31^. After removal of all APOE variants, combination of the two datasets resulted in a total of 85 independent SNPs (**Supplementary Table 1**; linkage disequilibrium [LD] r^2^≤0.2 in the European 1000G populations CEU, GBR, IBS, and TSI).

#### Generation of polygenic scores

PGS were calculated in PLINK 2.0 (www.cog-genomics.org/plink/2.0/) ^76^ as the sum over the weighted number of alleles per SNP, using the respective log(OR) as weights. Stage II log(OR) were used for SNPs from (Bellenguez et al., 2020). PGS were then normalized by the number of included variants (n=85) and finally standardized across all participants.

#### Generation of cell-type–specific genetic scores

Variants were included if their closest gene exceeded a relative expression level of 0.1 across 7 cell types: GABAergic neurons, glutamatergic neurons, microglia, oligodendrocytes, Oligodendrocyte Precursor Cells [OPC], astrocytes, endothelial ^77^. Cell types classified as “None” were excluded. Using Seurat 4.0.1 ^78^, we extracted expression levels from annotated human middle temporal gyrus (MTG) single-nucleus transcriptomes in the Allen Brain Atlas (15,928 total nuclei from 8 human tissue donors aged between 24 and 66 years)^79^. 25 principal components (PCs) were chosen for dimensionality reduction. rs7157106 and rs10131280 from the IGH cluster were excluded from the analysis.

### Tau-PET and amyloid-PET measurements

Tau-PET was assessed with a standardized protocol using 6 × 5 min frames, 75-105 min post-injection of [18F]AV1451. Similarly, amyloid-PET was acquired in 4 × 5 min frames, 50-70 min post injection of [18F]AV45 or 4 × 5 min frames, 90-110 min post injection of [18F]FBB. Recorded images were coregistered and averaged, and further standardized with respect to the orientation, voxel size and intensity by the ADNI PET core ^80^. Tau-PET scans were corrected for partial volume effects using the Geometric Transfer Matrix approach as previously described ^81^.

Tau-PET standardized uptake value ratio (SUVR) values were computed by normalizing the target regions of interest (ROIs) to the mean tracer uptake of the inferior cerebellar grey, following previous recommendations ^82,83^. Summary Tau-PET measures included average SUVR of ROIs defined by the Braak-staging: Braak stage 1 (entorhinal), Braak 3+4 (limbic) and Braak 5+6 (neocortical)^20^. We did not include Braak stage 2 (hippocampus) due to potential confounding signal from known off-target binding of the tau-PET tracer to the choroid plexus ^84^. The global tau-PET value was the mean of the FreeSurfer-derived cortical tau-PET SUVRs (excluding the hippocampus), computed for each subject.

The global amyloid-PET value was the mean FreeSurfer-derived cortical grey matter SUVRs (frontal, lateral temporal, lateral parietal and anterior/posterior cingulate) divided by whole cerebellum reference region, following previous recommendations ^85^. To obtain comparable quantification of the Aβ levels across tracers, we converted the global amyloid-PET values using the following centiloid calculation as recommended for the ADNI pipeline: AV45 centiloid = 196.9 × SUVR_FBP_ - 196.03, where SUVR_FBP_ is the SUVR of AV45, and FBB centiloid = 159.08 × SUVR_FBB_ – 151.65, where SUVR_FBB_ is the SUVR of FBB.

### Amyloid status

For each subject, the baseline global amyloid-PET values were binarized based on a cut-off of SUVR > 1.11 for global AV45-PET or SUVR > 1.08 for global FBB-PET to define the Aβ+ vs Aβ- status ^86^. For subjects without an available, the baseline CSF Aβ_1-42_ measurement was binarized, using the cut-off CSF Aβ_1-42_ < 976.6 pg/ml to define the to define the Aβ+ vs Aβ- status ^87^. Subjects with both CSF Aβ_1-42_ and amyloid-PET were considered positive if either one of the measures was abnormal.

### Cognitive assessment

Global cognition was assessed through the ADAS13, which is an extension of the 11-item cognitive subscale of the ADAS ^88^, including in addition a test of delayed word recall and number cancellation ^89^. Higher ADAS13 score means worse cognitive performance. Memory performance was assessed using the pre-established composite memory score ADNI-MEM ^90^. The ADNI-MEM score includes the Rey Auditory Verbal Learning Test, the Alzheimer’s Disease Assessment Scale, the Wechsler Logical Memory I and II, and the word recall of the MMSE ^90^.

### Statistical analysis

To ensure that our findings were not affected by outliers, biomarker measurements deviating ±3 standard deviations from the sample mean were excluded. Including outliers in a sensitivity analysis yielded consistent results. Tau-PET SUVR measures were log-transformed prior to analysis to approximate a normal distribution.

In order to examine the associations between the PGS and the rates of change in either tau-PET, amyloid-PET or cognition, we first determined the subject-level rate of change for each of the biomarkers and cognitive performance, using a previously established approach ^91^. To that end, we employed linear mixed-effect regression analyses to model the rate of change at the subject-level, including time as the independent variable, with the random terms being slope and intercept. Using the thus estimated rates of change as the dependent variables, we tested in univariate linear regression analyses the PGS as the predictor, controlled for age, sex, education, diagnosis, ethnicity, APOE genotype, the first ten principal components to correct for population stratification and maximum follow-up time. Similarly, for cross-sectional data, we tested in separate regression analyses whether the PGS predicted the different biomarkers, adjusting for age, sex, years of education, diagnosis, self-reported ethnicity, APOE genotype, and the first ten principal components.

To test whether the PGS effect on cognitive changes was mediated by changes in tau-PET, we conducted mediation analyses, testing whether the associations between PGS and changes in cognition were mediated via change in global tau-PET uptake. Significance of the mediation was assed using 1,000 bootstrapped iterations, as implemented in “mediation” package in R. Mediation model was controlled for age, sex, education, diagnosis, ethnicity, APOE genotype and first ten principal components.

Next, we performed sensitivity analyses in subgroups categorized by amyloid status, where the effect of PGS on tau-PET change was tested in each of the subgroups. Significance and 95%- confidence intervals (CI) of effects were determined using 1000 bootstrapping iterations. We further tested the interaction between the PGS and global amyloid-PET centiloid on tau-PET change rate, controlling for age, sex, education, diagnosis, ethnicity, APOE genotype, the first ten principal components and maximum follow-up time.

Finally, we estimated sample size required for detection hypothetical treatment effects on the rate of change in tau-PET at power = 0.8 and treatment effect size of 20% or 40%. In a first, step, the subject-level PGSs were residualized by regressing out age, sex, education, diagnosis, ethnicity, APOE genotype, and the first ten principal components in order to render the stratified analyses of the PGS - to be conducted in the next step - independent of these potential confounds. Subjects were stratified into quartiles of the residualized PGSs and sample size estimates were conducted using the “pwr” R package using the following parameters: two-sample t-test, two-tailed, type I error rate = 0.05, power = 0.8. Sample size estimates were repeated for alternative stratifications including Aβ status (Aβ+ vs Aβ+), APOE ε4 status (ε4 carrier vs non-carrier), and combinations of these stratification factors.

All statistical analyses were performed using R (http://www.R-project.org). Results were corrected for multiple comparisons using the false discovery rate (FDR) ^92^ with a significance level of 0.05. Standardized beta-coefficients are reported throughout to facilitate comparison of associations across biomarkers.

## Data availability statement

The data that used in this study were obtained from the Alzheimer’s disease Neuroimaging Initiative (ADNI) and are available from the ADNI database (adni.loni.usc.edu) upon registration and compliance with the data usage agreement. A source file for all figures as can be found in the supplementary.

## Data Availability

The data that used in this study were obtained from the Alzheimer's disease Neuroimaging Initiative (ADNI) and are available from the ADNI database (adni.loni.usc.edu) upon registration and compliance with the data usage agreement.

## Author contributions

A.R. data processing, writing the manuscript

S.F. data processing, critical revision of the manuscript

N.F. critical revision of the manuscript

R.M. data processing, critical revision of the manuscript

A.R. critical revision of the manuscript

M.D. critical revision of the manuscript

M.E. study concept and design, interpretation of the results, writing the manuscript

ADNI provided data used for this study

## Acknowledgements

Parts of the data used in preparation of this manuscript were obtained from the ADNI database (adni.loni.usc.edu). As such, the investigators within the ADNI study contributed to the design and implementation of ADNI and/or provided data but did not participate in analysis or writing of this paper. A complete listing of ADNI investigators can be found at the end of the manuscript.

