## Supplementary figures for "Polygenic effect on accelerated tau pathology accumulation in Alzheimer’s disease: implications for patient selection in clinical trials"

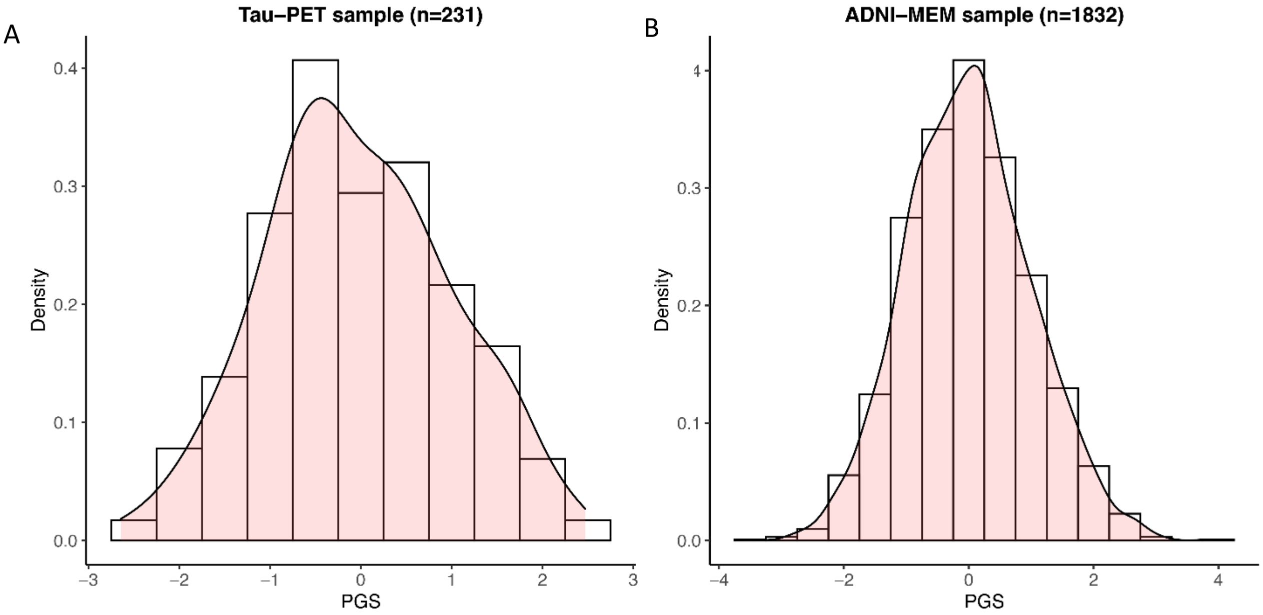


**Supplementary Figure 1:** PGS distribution within the tau-PET sample **(A)** and the larger ADNI-MEM sample **(B)**.

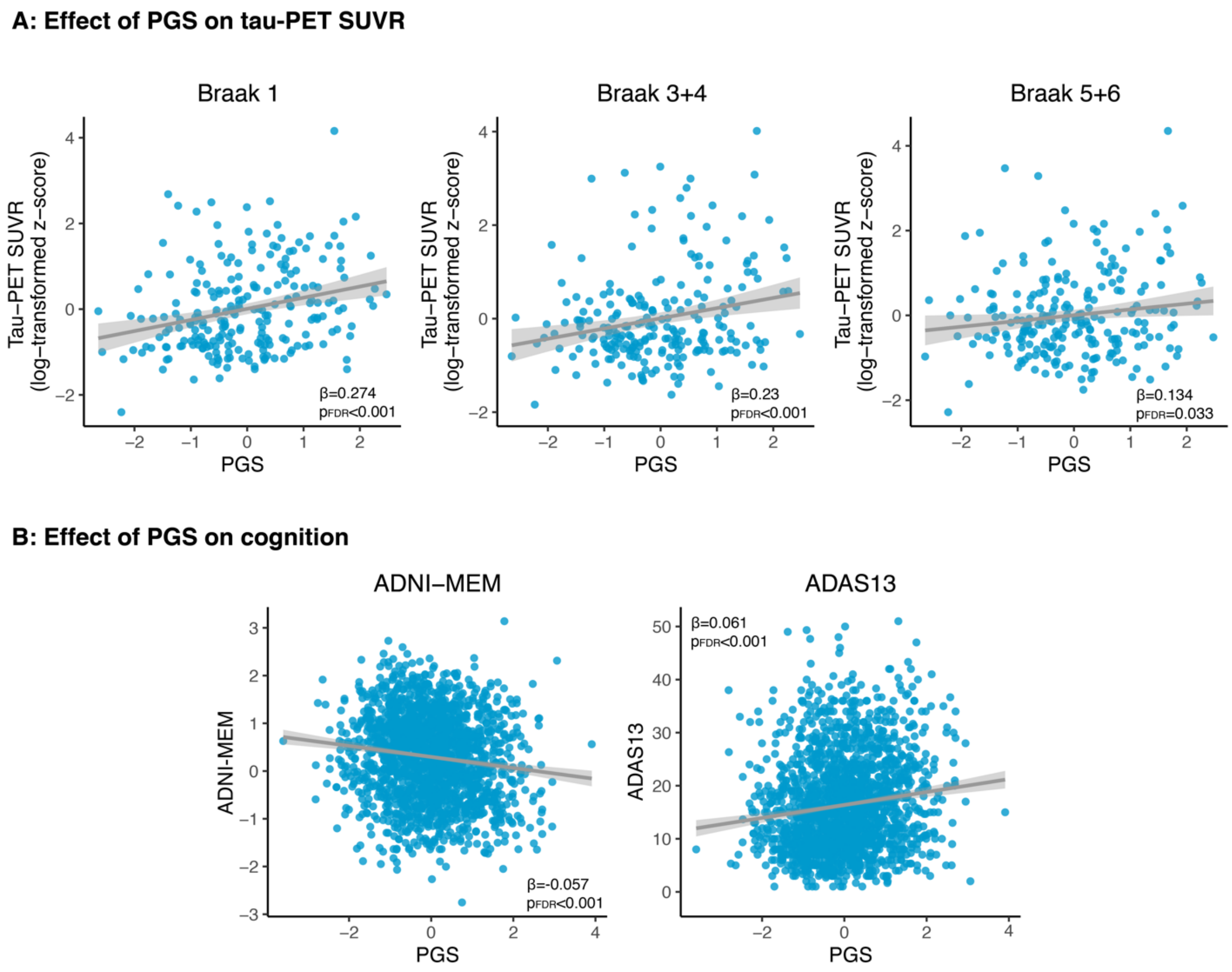


**Supplementary Figure 2:** Scatterplots representing the significant associations between PGS and cross-sectional tau **(A)** and cognition **(B)**.
