## Supplementary tables for "Polygenic effect on accelerated tau pathology accumulation in Alzheimer’s disease: implications for patient selection in clinical trials"

**Supplementary Table 1:** List of SNPs included in the analysis

| rsid | Gene | chr | pos_hg19 | A1 | A2 | A1freq | b | OR* | p | source | novel |
|---|---|---|---|---|---|---|---|---|---|---|---|
| rs141749679 | SORT1 | 1 | 109888432 | C | T | 0.004 | 0.344 | 1.41 | 7.54E-09 | Bellenguez | 1 |
| rs679515 | CR1 | 1 | 207750568 | T | C | 0.188 | 0.113 | 1.12 | 7.16E-46 | Bellenguez | 0 |
| rs113020870 | AGRN | 1 | 985377 | T | C | 0.004 | 0.069 | 1.07 | 3.83E-08 | Wightman | 1 |
| rs143080277 | NCK2 | 2 | 106366056 | C | T | 0.005 | 0.365 | 1.44 | 2.07E-13 | Bellenguez | 1 |
| rs6733839 | BIN1 | 2 | 127892810 | T | C | 0.389 | 0.148 | 1.16 | 6.06E-118 | Bellenguez | 0 |
| rs10933431 | INPP5D | 2 | 233981912 | C | G | 0.766 | 0.041 | 1.04 | 3.62E-18 | Bellenguez | 0 |
| rs17020490 | PRKD3 | 2 | 37531939 | C | T | 0.145 | 0.058 | 1.06 | 3.29E-09 | Bellenguez | 1 |
| rs72777026 | ADAM17 | 2 | 9699011 | G | A | 0.144 | 0.058 | 1.06 | 2.72E-08 | Bellenguez | 1 |
| rs7597763 | INPP5D | 2 | 234082577 | C | A | 0.45 | 0.009 | 1.01 | 4.65E-09 | Wightman | 0 |
| rs16824536 | MME | 3 | 154787511 | G | A | 0.946 | 0.083 | 1.09 | 3.63E-08 | Bellenguez | 1 |
| rs61762319 | MME | 3 | 154801978 | G | A | 0.026 | 0.166 | 1.18 | 2.16E-11 | Bellenguez | 1 |
| rs6846529 | HS3ST1 | 4 | 11025131 | C | T | 0.283 | 0.058 | 1.06 | 2.20E-17 | Bellenguez | 0 |
| rs2245466 | RHOH | 4 | 40198846 | G | C | 0.343 | 0.049 | 1.05 | 1.22E-09 | Bellenguez | 1 |
| rs3822030 | IDUA | 4 | 987343 | T | G | 0.571 | 0.041 | 1.04 | 8.29E-12 | Bellenguez | 1 |
| rs112403360 | ANKH | 5 | 14724413 | A | T | 0.073 | 0.131 | 1.14 | 2.27E-09 | Bellenguez | 1 |
| rs871269 | TNIP1 | 5 | 150432388 | C | T | 0.674 | 0.041 | 1.04 | 8.67E-09 | Bellenguez | 1 |
| rs113706587 | RASGEF1C | 5 | 179628150 | A | G | 0.11 | 0.086 | 1.09 | 2.22E-16 | Bellenguez | 1 |
| rs62374257 | COX7C | 5 | 86223195 | C | T | 0.23 | 0.058 | 1.06 | 1.38E-15 | Bellenguez | 1 |
| rs6891966 | HAVCR2 | 5 | 156526331 | G | A | 0.77 | 0.01 | 1.01 | 7.91E-10 | Wightman | 1 |
| rs785129 | HS3ST5 | 6 | 114612895 | T | C | 0.35 | 0.039 | 1.04 | 2.40E-09 | Bellenguez | 1 |
| rs6605556 | HLA-DQA1 | 6 | 32583099 | A | G | 0.839 | 0.062 | 1.06 | 7.07E-20 | Bellenguez | 0 |
| rs10947943 | UNC5CL | 6 | 41004093 | G | A | 0.858 | 0.073 | 1.08 | 1.13E-09 | Bellenguez | 0 |
| rs143332484 | TREM2 | 6 | 41129207 | T | C | 0.013 | 0.365 | 1.44 | 2.78E-25 | Bellenguez | 0 |
| rs75932628 | TREM2 | 6 | 41129252 | T | C | 0.003 | 0.833 | 2.3 | 2.53E-37 | Bellenguez | 0 |
| rs60755019 | TREML2 | 6 | 41149008 | G | A | 0.004 | 0.412 | 1.51 | 2.07E-08 | Bellenguez | 0 |
| rs7767350 | CD2AP | 6 | 47485126 | T | C | 0.271 | 0.095 | 1.1 | 7.94E-22 | Bellenguez | 0 |
| rs13237518 | TMEM106B | 7 | 12269593 | C | A | 0.588 | 0.051 | 1.05 | 4.88E-11 | Bellenguez | 1 |
| rs11771145 | EPHA1 | 7 | 143110762 | G | A | 0.652 | 0.041 | 1.04 | 3.30E-14 | Bellenguez | 0 |
| rs6966331 | EPDR1 | 7 | 37883793 | C | T | 0.651 | 0.051 | 1.05 | 4.64E-10 | Bellenguez | 0 |
| rs76928645 | SEC61G | 7 | 54941328 | C | T | 0.897 | 0.062 | 1.06 | 1.62E-10 | Bellenguez | 1 |
| rs6943429 | UMAD1 | 7 | 7856894 | T | C | 0.421 | 0.049 | 1.05 | 1.03E-10 | Bellenguez | 1 |
| rs10952097 | ICA1 | 7 | 8244012 | T | C | 0.114 | 0.068 | 1.07 | 6.81E-09 | Bellenguez | 1 |

| | | | | | | | | | | | |
|---|---|---|---|---|---|---|---|---|---|---|---|
| rs7384878 | SPDYE3 | 7 | 99932049 | T | C | 0.69 | 0.083 | 1.09 | 1.06E-26 | Bellenguez | 0 |
| rs1065712 | CTSB | 8 | 11702122 | C | G | 0.053 | 0.058 | 1.06 | 1.94E-09 | Bellenguez | 1 |
| rs34173062 | SHARPIN | 8 | 145158607 | A | G | 0.081 | 0.131 | 1.14 | 1.72E-16 | Bellenguez | 1 |
| rs73223431 | PTK2B | 8 | 27219987 | T | C | 0.369 | 0.068 | 1.07 | 4.03E-22 | Bellenguez | 0 |
| rs11787077 | CLU | 8 | 27465312 | C | T | 0.608 | 0.083 | 1.09 | 1.70E-44 | Bellenguez | 0 |
| rs61732533 | OPLAH | 8 | 145108151 | A | G | 0.05 | 0.018 | 1.02 | 3.14E-09 | Wightman | 0 |
| rs1800978 | ABCA1 | 9 | 107665978 | G | C | 0.13 | 0.039 | 1.04 | 1.59E-09 | Bellenguez | 1 |
| rs7912495 | USP6NL | 10 | 11718713 | G | A | 0.462 | 0.068 | 1.07 | 9.74E-19 | Bellenguez | 0 |
| rs7908662 | PLEKHA1 | 10 | 124172912 | A | G | 0.533 | 0.041 | 1.04 | 2.59E-09 | Bellenguez | 1 |
| rs7068231 | ANK3 | 10 | 61784928 | G | T | 0.597 | 0.051 | 1.05 | 3.32E-13 | Bellenguez | 1 |
| rs6586028 | TSPAN14 | 10 | 82253984 | T | C | 0.804 | 0.073 | 1.08 | 1.97E-19 | Bellenguez | 1 |
| rs6584063 | BLNK | 10 | 98026407 | A | G | 0.956 | 0.128 | 1.14 | 6.73E-11 | Bellenguez | 1 |
| rs74685827 | SORL1 | 11 | 121353077 | G | T | 0.019 | 0.104 | 1.11 | 2.81E-11 | Bellenguez | 0 |
| rs11218343 | SORL1 | 11 | 121435587 | T | C | 0.961 | 0.198 | 1.22 | 1.40E-21 | Bellenguez | 0 |
| rs10437655 | SPI1 | 11 | 47391948 | A | G | 0.399 | 0.049 | 1.05 | 5.28E-14 | Bellenguez | 0 |
| rs1582763 | MS4A4A | 11 | 60021948 | G | A | 0.629 | 0.117 | 1.12 | 3.74E-42 | Bellenguez | 0 |
| rs3851179 | PICALM | 11 | 85868640 | C | T | 0.642 | 0.094 | 1.1 | 2.95E-48 | Bellenguez | 0 |
| rs6489896 | TPCN1 | 12 | 113719788 | C | T | 0.076 | 0.077 | 1.08 | 1.80E-09 | Bellenguez | 1 |
| rs7157106 | IGHG1 | 14 | 106228095 | A | G | 0.361 | 0.039 | 1.04 | 1.99E-08 | Bellenguez | 1 |
| rs10131280 | IGH_cluster | 14 | 107121607 | G | A | 0.867 | 0.062 | 1.06 | 4.26E-10 | Bellenguez | 1 |
| rs17125924 | FERMT2 | 14 | 53391680 | G | A | 0.089 | 0.104 | 1.11 | 8.32E-16 | Bellenguez | 0 |
| rs7401792 | SLC24A4 | 14 | 92931261 | G | A | 0.371 | 0.039 | 1.04 | 4.83E-08 | Bellenguez | 0 |
| rs12590654 | SLC24A4 | 14 | 92938855 | G | A | 0.672 | 0.062 | 1.06 | 4.25E-21 | Bellenguez | 0 |
| rs8025980 | SPPL2A | 15 | 50994011 | A | G | 0.655 | 0.062 | 1.06 | 1.32E-08 | Bellenguez | 0 |
| rs602602 | MINDY2 | 15 | 59057023 | T | A | 0.72 | 0.051 | 1.05 | 2.07E-15 | Bellenguez | 0 |
| rs117618017 | APH1B | 15 | 63569902 | T | C | 0.144 | 0.086 | 1.09 | 2.15E-25 | Bellenguez | 0 |
| rs3848143 | SNX1 | 15 | 64423506 | G | A | 0.22 | 0.068 | 1.07 | 8.41E-11 | Bellenguez | 1 |
| rs12592898 | CTSH | 15 | 79229199 | G | A | 0.867 | 0.062 | 1.06 | 4.18E-09 | Bellenguez | 1 |
| rs889555 | BCKDK | 16 | 31122571 | C | T | 0.719 | 0.041 | 1.04 | 1.96E-11 | Bellenguez | 0 |
| rs4985556 | IL34 | 16 | 70694000 | A | C | 0.115 | 0.086 | 1.09 | 5.98E-10 | Bellenguez | 0 |
| rs450674 | MAF | 16 | 79608408 | T | C | 0.627 | 0.02 | 1.02 | 3.16E-08 | Bellenguez | 1 |
| rs12446759 | PLCG2 | 16 | 81773003 | A | G | 0.597 | 0.041 | 1.04 | 1.22E-13 | Bellenguez | 0 |
| rs72824905 | PLCG2 | 16 | 81942028 | C | G | 0.992 | 0.248 | 1.28 | 8.48E-12 | Bellenguez | 0 |
| rs16941239 | FOXF1 | 16 | 86454210 | A | T | 0.029 | 0.113 | 1.12 | 1.29E-08 | Bellenguez | 1 |
| rs56407236 | PRDM7 | 16 | 90170095 | A | G | 0.069 | 0.104 | 1.11 | 6.47E-15 | Bellenguez | 1 |

| | | | | | | | | | | | |
|---|---|---|---|---|---|---|---|---|---|---|---|
| rs2242595 | MYO15A | 17 | 18059454 | G | A | 0.888 | 0.073 | 1.08 | 1.11E-09 | Bellenguez | 1 |
| rs5848 | GRN | 17 | 42430244 | T | C | 0.289 | 0.077 | 1.08 | 2.38E-20 | Bellenguez | 1 |
| rs199515 | WNT3 | 17 | 44856641 | C | G | 0.781 | 0.073 | 1.08 | 9.34E-13 | Bellenguez | 0 |
| rs7225151 | SCIMP | 17 | 5137047 | A | G | 0.124 | 0.049 | 1.05 | 4.13E-13 | Bellenguez | 0 |
| rs2526377 | TSPOAP1 | 17 | 56410041 | A | G | 0.555 | 0.051 | 1.05 | 1.58E-12 | Bellenguez | 0 |
| rs4277405 | ACE | 17 | 61548918 | T | C | 0.616 | 0.051 | 1.05 | 8.80E-20 | Bellenguez | 0 |
| rs28394864 | ZNF652 | 17 | 47450775 | A | G | 0.46 | 0.009 | 1.01 | 4.90E-10 | Wightman | 0 |
| rs12151021 | ABCA7 | 19 | 1050874 | A | G | 0.336 | 0.086 | 1.09 | 1.59E-37 | Bellenguez | 0 |
| rs9304690 | SIGLEC11 | 19 | 50453317 | T | C | 0.24 | 0.058 | 1.06 | 4.74E-09 | Bellenguez | 1 |
| rs587709 | LILRB2 | 19 | 54771451 | C | T | 0.325 | 0.049 | 1.05 | 3.63E-11 | Bellenguez | 1 |
| rs2452170 | MAMSTR | 19 | 49213504 | A | G | 0.53 | 0.008 | 1.01 | 1.72E-08 | Wightman | 1 |
| rs1354106 | CD33 | 19 | 51737991 | T | G | 0.63 | 0.011 | 1.01 | 2.21E-10 | Wightman | 0 |
| rs1761461 | LILRA5 | 19 | 54825174 | C | A | 0.49 | 0.008 | 1.01 | 1.56E-09 | Wightman | 0 |
| rs1358782 | RBCK1 | 20 | 393978 | G | A | 0.754 | 0.041 | 1.04 | 1.55E-08 | Bellenguez | 1 |
| rs6014724 | CASS4 | 20 | 54998544 | A | G | 0.91 | 0.117 | 1.12 | 4.13E-21 | Bellenguez | 0 |
| rs6742 | SLC2A4RG | 20 | 62374441 | C | T | 0.779 | 0.062 | 1.06 | 2.58E-09 | Bellenguez | 1 |
| rs2154481 | APP | 21 | 27473875 | T | C | 0.524 | 0.041 | 1.04 | 1.00E-12 | Bellenguez | 1 |
| rs2830489 | ADAMTS1 | 21 | 28148191 | C | T | 0.719 | 0.03 | 1.03 | 1.69E-10 | Bellenguez | 0 |

rsid = rs number; Gene = nearest protein-coding gene; chr = chromosome; pos_hg19 = position (hg19); A1 = effect allele; A2 = other allele; A1freq = A1 frequency; b = beta; OR = odds ratio referring to A1; p = p-value; source = source GWAS; novel = novel SNP
*OR based on the Stage II analysis for SNPs from Bellenguez et al.

**Supplementary Table 2:** List of SNPs included in the cell-type-specific scores

| rsid | Gene | chr | pos_hg19 | A1 | A2 | A1freq | b | OR* | p | source | Celltype | Expression |
|---|---|---|---|---|---|---|---|---|---|---|---|---|
| rs7912495 | USP6NL | 10 | 11718713 | G | A | 0.462 | 0.068 | 1.07 | 9.74E-19 | Bellenguez | Astrocytes | 0.028380657 |
| rs7912495 | USP6NL | 10 | 11718713 | G | A | 0.462 | 0.068 | 1.07 | 9.74E-19 | Bellenguez | Endothelial | 0.355104284 |
| rs7912495 | USP6NL | 10 | 11718713 | G | A | 0.462 | 0.068 | 1.07 | 9.74E-19 | Bellenguez | GABAergic | 0.072374124 |
| rs7912495 | USP6NL | 10 | 11718713 | G | A | 0.462 | 0.068 | 1.07 | 9.74E-19 | Bellenguez | Glutamatergic | 0.053938237 |
| rs7912495 | USP6NL | 10 | 11718713 | G | A | 0.462 | 0.068 | 1.07 | 9.74E-19 | Bellenguez | Microglia | 0.332558185 |
| rs7912495 | USP6NL | 10 | 11718713 | G | A | 0.462 | 0.068 | 1.07 | 9.74E-19 | Bellenguez | Oligodendrocytes | 0.029791626 |
| rs7912495 | USP6NL | 10 | 11718713 | G | A | 0.462 | 0.068 | 1.07 | 9.74E-19 | Bellenguez | OPC | 0.127852887 |
| rs7908662 | PLEKHA1 | 10 | 124172912 | A | G | 0.533 | 0.041 | 1.04 | 2.59E-09 | Bellenguez | Astrocytes | 0.043037072 |
| rs7908662 | PLEKHA1 | 10 | 124172912 | A | G | 0.533 | 0.041 | 1.04 | 2.59E-09 | Bellenguez | Endothelial | 0.015532954 |
| rs7908662 | PLEKHA1 | 10 | 124172912 | A | G | 0.533 | 0.041 | 1.04 | 2.59E-09 | Bellenguez | GABAergic | 0.207969672 |
| rs7908662 | PLEKHA1 | 10 | 124172912 | A | G | 0.533 | 0.041 | 1.04 | 2.59E-09 | Bellenguez | Glutamatergic | 0.307975082 |
| rs7908662 | PLEKHA1 | 10 | 124172912 | A | G | 0.533 | 0.041 | 1.04 | 2.59E-09 | Bellenguez | Microglia | 0.042029274 |
| rs7908662 | PLEKHA1 | 10 | 124172912 | A | G | 0.533 | 0.041 | 1.04 | 2.59E-09 | Bellenguez | Oligodendrocytes | 0.320042911 |
| rs7908662 | PLEKHA1 | 10 | 124172912 | A | G | 0.533 | 0.041 | 1.04 | 2.59E-09 | Bellenguez | OPC | 0.063413035 |
| rs7068231 | ANK3 | 10 | 61784928 | G | T | 0.597 | 0.051 | 1.05 | 3.32E-13 | Bellenguez | Astrocytes | 0.00374241 |
| rs7068231 | ANK3 | 10 | 61784928 | G | T | 0.597 | 0.051 | 1.05 | 3.32E-13 | Bellenguez | Endothelial | 0.000148053 |
| rs7068231 | ANK3 | 10 | 61784928 | G | T | 0.597 | 0.051 | 1.05 | 3.32E-13 | Bellenguez | GABAergic | 0.216784838 |
| rs7068231 | ANK3 | 10 | 61784928 | G | T | 0.597 | 0.051 | 1.05 | 3.32E-13 | Bellenguez | Glutamatergic | 0.18576997 |
| rs7068231 | ANK3 | 10 | 61784928 | G | T | 0.597 | 0.051 | 1.05 | 3.32E-13 | Bellenguez | Microglia | 0.050372863 |
| rs7068231 | ANK3 | 10 | 61784928 | G | T | 0.597 | 0.051 | 1.05 | 3.32E-13 | Bellenguez | Oligodendrocytes | 0.408262837 |
| rs7068231 | ANK3 | 10 | 61784928 | G | T | 0.597 | 0.051 | 1.05 | 3.32E-13 | Bellenguez | OPC | 0.13491903 |
| rs6586028 | TSPAN14 | 10 | 82253984 | T | C | 0.804 | 0.073 | 1.08 | 1.97E-19 | Bellenguez | Astrocytes | 0.088346796 |
| rs6586028 | TSPAN14 | 10 | 82253984 | T | C | 0.804 | 0.073 | 1.08 | 1.97E-19 | Bellenguez | Endothelial | 0.105974466 |
| rs6586028 | TSPAN14 | 10 | 82253984 | T | C | 0.804 | 0.073 | 1.08 | 1.97E-19 | Bellenguez | GABAergic | 0.085070071 |
| rs6586028 | TSPAN14 | 10 | 82253984 | T | C | 0.804 | 0.073 | 1.08 | 1.97E-19 | Bellenguez | Glutamatergic | 0.109517552 |
| rs6586028 | TSPAN14 | 10 | 82253984 | T | C | 0.804 | 0.073 | 1.08 | 1.97E-19 | Bellenguez | Microglia | 0.43654748 |
| rs6586028 | TSPAN14 | 10 | 82253984 | T | C | 0.804 | 0.073 | 1.08 | 1.97E-19 | Bellenguez | Oligodendrocytes | 0.107829344 |
| rs6586028 | TSPAN14 | 10 | 82253984 | T | C | 0.804 | 0.073 | 1.08 | 1.97E-19 | Bellenguez | OPC | 0.066714291 |
| rs6584063 | BLNK | 10 | 98026407 | A | G | 0.956 | 0.128 | 1.14 | 6.73E-11 | Bellenguez | Astrocytes | 0.00916639 |
| rs6584063 | BLNK | 10 | 98026407 | A | G | 0.956 | 0.128 | 1.14 | 6.73E-11 | Bellenguez | Endothelial | 0 |

| | | | | | | | | | | | | |
|---|---|---|---|---|---|---|---|---|---|---|---|---|
| rs6584063 | BLNK | 10 | 98026407 | A | G | 0.956 | 0.128 | 1.14 | 6.73E-11 | Bellenguez | GABAergic | 0.00328515 |
| rs6584063 | BLNK | 10 | 98026407 | A | G | 0.956 | 0.128 | 1.14 | 6.73E-11 | Bellenguez | Glutamatergic | 0.003969826 |
| rs6584063 | BLNK | 10 | 98026407 | A | G | 0.956 | 0.128 | 1.14 | 6.73E-11 | Bellenguez | Microglia | 0.960212585 |
| rs6584063 | BLNK | 10 | 98026407 | A | G | 0.956 | 0.128 | 1.14 | 6.73E-11 | Bellenguez | Oligodendrocytes | 0.017942151 |
| rs6584063 | BLNK | 10 | 98026407 | A | G | 0.956 | 0.128 | 1.14 | 6.73E-11 | Bellenguez | OPC | 0.005423899 |
| rs74685827 | SORL1 | 11 | 121353077 | G | T | 0.019 | 0.104 | 1.11 | 2.81E-11 | Bellenguez | Astrocytes | 0.087980118 |
| rs74685827 | SORL1 | 11 | 121353077 | G | T | 0.019 | 0.104 | 1.11 | 2.81E-11 | Bellenguez | Endothelial | 0 |
| rs74685827 | SORL1 | 11 | 121353077 | G | T | 0.019 | 0.104 | 1.11 | 2.81E-11 | Bellenguez | GABAergic | 0.043761186 |
| rs74685827 | SORL1 | 11 | 121353077 | G | T | 0.019 | 0.104 | 1.11 | 2.81E-11 | Bellenguez | Glutamatergic | 0.096739051 |
| rs74685827 | SORL1 | 11 | 121353077 | G | T | 0.019 | 0.104 | 1.11 | 2.81E-11 | Bellenguez | Microglia | 0.669377459 |
| rs74685827 | SORL1 | 11 | 121353077 | G | T | 0.019 | 0.104 | 1.11 | 2.81E-11 | Bellenguez | Oligodendrocytes | 0.034385347 |
| rs74685827 | SORL1 | 11 | 121353077 | G | T | 0.019 | 0.104 | 1.11 | 2.81E-11 | Bellenguez | OPC | 0.067756838 |
| rs11218343 | SORL1 | 11 | 121435587 | T | C | 0.961 | 0.198 | 1.22 | 1.4E-21 | Bellenguez | Astrocytes | 0.087980118 |
| rs11218343 | SORL1 | 11 | 121435587 | T | C | 0.961 | 0.198 | 1.22 | 1.4E-21 | Bellenguez | Endothelial | 0 |
| rs11218343 | SORL1 | 11 | 121435587 | T | C | 0.961 | 0.198 | 1.22 | 1.4E-21 | Bellenguez | GABAergic | 0.043761186 |
| rs11218343 | SORL1 | 11 | 121435587 | T | C | 0.961 | 0.198 | 1.22 | 1.4E-21 | Bellenguez | Glutamatergic | 0.096739051 |
| rs11218343 | SORL1 | 11 | 121435587 | T | C | 0.961 | 0.198 | 1.22 | 1.4E-21 | Bellenguez | Microglia | 0.669377459 |
| rs11218343 | SORL1 | 11 | 121435587 | T | C | 0.961 | 0.198 | 1.22 | 1.4E-21 | Bellenguez | Oligodendrocytes | 0.034385347 |
| rs11218343 | SORL1 | 11 | 121435587 | T | C | 0.961 | 0.198 | 1.22 | 1.4E-21 | Bellenguez | OPC | 0.067756838 |
| rs10437655 | SPI1 | 11 | 47391948 | A | G | 0.399 | 0.049 | 1.05 | 5.28E-14 | Bellenguez | Astrocytes | 0.131847623 |
| rs10437655 | SPI1 | 11 | 47391948 | A | G | 0.399 | 0.049 | 1.05 | 5.28E-14 | Bellenguez | Endothelial | 0 |
| rs10437655 | SPI1 | 11 | 47391948 | A | G | 0.399 | 0.049 | 1.05 | 5.28E-14 | Bellenguez | GABAergic | 0.044167387 |
| rs10437655 | SPI1 | 11 | 47391948 | A | G | 0.399 | 0.049 | 1.05 | 5.28E-14 | Bellenguez | Glutamatergic | 0.024467464 |
| rs10437655 | SPI1 | 11 | 47391948 | A | G | 0.399 | 0.049 | 1.05 | 5.28E-14 | Bellenguez | Microglia | 0.737250317 |
| rs10437655 | SPI1 | 11 | 47391948 | A | G | 0.399 | 0.049 | 1.05 | 5.28E-14 | Bellenguez | Oligodendrocytes | 0.011783551 |
| rs10437655 | SPI1 | 11 | 47391948 | A | G | 0.399 | 0.049 | 1.05 | 5.28E-14 | Bellenguez | OPC | 0.050483657 |
| rs1582763 | MS4A4A | 11 | 60021948 | G | A | 0.629 | 0.117 | 1.12 | 3.74E-42 | Bellenguez | Astrocytes | 0 |
| rs1582763 | MS4A4A | 11 | 60021948 | G | A | 0.629 | 0.117 | 1.12 | 3.74E-42 | Bellenguez | Endothelial | 0.002030941 |
| rs1582763 | MS4A4A | 11 | 60021948 | G | A | 0.629 | 0.117 | 1.12 | 3.74E-42 | Bellenguez | GABAergic | 2.03154E-05 |
| rs1582763 | MS4A4A | 11 | 60021948 | G | A | 0.629 | 0.117 | 1.12 | 3.74E-42 | Bellenguez | Glutamatergic | 0.001983229 |
| rs1582763 | MS4A4A | 11 | 60021948 | G | A | 0.629 | 0.117 | 1.12 | 3.74E-42 | Bellenguez | Microglia | 0.994931518 |
| rs1582763 | MS4A4A | 11 | 60021948 | G | A | 0.629 | 0.117 | 1.12 | 3.74E-42 | Bellenguez | Oligodendrocytes | 0 |
| rs1582763 | MS4A4A | 11 | 60021948 | G | A | 0.629 | 0.117 | 1.12 | 3.74E-42 | Bellenguez | OPC | 0.001033996 |
| rs3851179 | PICALM | 11 | 85868640 | C | T | 0.642 | 0.094 | 1.10 | 2.95E-48 | Bellenguez | Astrocytes | 0.060851449 |
| rs3851179 | PICALM | 11 | 85868640 | C | T | 0.642 | 0.094 | 1.10 | 2.95E-48 | Bellenguez | Endothelial | 0.241068222 |

| | | | | | | | | | | | | |
|---|---|---|---|---|---|---|---|---|---|---|---|---|
| rs3851179 | PICALM | 11 | 85868640 | C | T | 0.642 | 0.094 | 1.10 | 2.95E-48 | Bellenguez | GABAergic | 0.057088605 |
| rs3851179 | PICALM | 11 | 85868640 | C | T | 0.642 | 0.094 | 1.10 | 2.95E-48 | Bellenguez | Glutamatergic | 0.064033322 |
| rs3851179 | PICALM | 11 | 85868640 | C | T | 0.642 | 0.094 | 1.10 | 2.95E-48 | Bellenguez | Microglia | 0.217808499 |
| rs3851179 | PICALM | 11 | 85868640 | C | T | 0.642 | 0.094 | 1.10 | 2.95E-48 | Bellenguez | Oligodendrocytes | 0.293070234 |
| rs3851179 | PICALM | 11 | 85868640 | C | T | 0.642 | 0.094 | 1.10 | 2.95E-48 | Bellenguez | OPC | 0.066079668 |
| rs6489896 | TPCN1 | 12 | 113719788 | C | T | 0.076 | 0.077 | 1.08 | 1.8E-09 | Bellenguez | Astrocytes | 0.355311833 |
| rs6489896 | TPCN1 | 12 | 113719788 | C | T | 0.076 | 0.077 | 1.08 | 1.8E-09 | Bellenguez | Endothelial | 0.277098503 |
| rs6489896 | TPCN1 | 12 | 113719788 | C | T | 0.076 | 0.077 | 1.08 | 1.8E-09 | Bellenguez | GABAergic | 0.028541719 |
| rs6489896 | TPCN1 | 12 | 113719788 | C | T | 0.076 | 0.077 | 1.08 | 1.8E-09 | Bellenguez | Glutamatergic | 0.0276378 |
| rs6489896 | TPCN1 | 12 | 113719788 | C | T | 0.076 | 0.077 | 1.08 | 1.8E-09 | Bellenguez | Microglia | 0.169466457 |
| rs6489896 | TPCN1 | 12 | 113719788 | C | T | 0.076 | 0.077 | 1.08 | 1.8E-09 | Bellenguez | Oligodendrocytes | 0.08273171 |
| rs6489896 | TPCN1 | 12 | 113719788 | C | T | 0.076 | 0.077 | 1.08 | 1.8E-09 | Bellenguez | OPC | 0.059211979 |
| rs17125924 | FERMT2 | 14 | 53391680 | G | A | 0.089 | 0.104 | 1.11 | 8.32E-16 | Bellenguez | Astrocytes | 0.306081714 |
| rs17125924 | FERMT2 | 14 | 53391680 | G | A | 0.089 | 0.104 | 1.11 | 8.32E-16 | Bellenguez | Endothelial | 0.316384241 |
| rs17125924 | FERMT2 | 14 | 53391680 | G | A | 0.089 | 0.104 | 1.11 | 8.32E-16 | Bellenguez | GABAergic | 0.033631604 |
| rs17125924 | FERMT2 | 14 | 53391680 | G | A | 0.089 | 0.104 | 1.11 | 8.32E-16 | Bellenguez | Glutamatergic | 0.032605899 |
| rs17125924 | FERMT2 | 14 | 53391680 | G | A | 0.089 | 0.104 | 1.11 | 8.32E-16 | Bellenguez | Microglia | 0.018858734 |
| rs17125924 | FERMT2 | 14 | 53391680 | G | A | 0.089 | 0.104 | 1.11 | 8.32E-16 | Bellenguez | Oligodendrocytes | 0.149864908 |
| rs17125924 | FERMT2 | 14 | 53391680 | G | A | 0.089 | 0.104 | 1.11 | 8.32E-16 | Bellenguez | OPC | 0.1425729 |
| rs7401792 | SLC24A4 | 14 | 92931261 | G | A | 0.371 | 0.039 | 1.04 | 4.83E-08 | Bellenguez | Astrocytes | 0.156636464 |
| rs7401792 | SLC24A4 | 14 | 92931261 | G | A | 0.371 | 0.039 | 1.04 | 4.83E-08 | Bellenguez | Endothelial | 0 |
| rs7401792 | SLC24A4 | 14 | 92931261 | G | A | 0.371 | 0.039 | 1.04 | 4.83E-08 | Bellenguez | GABAergic | 0.337108325 |
| rs7401792 | SLC24A4 | 14 | 92931261 | G | A | 0.371 | 0.039 | 1.04 | 4.83E-08 | Bellenguez | Glutamatergic | 0.283856164 |
| rs7401792 | SLC24A4 | 14 | 92931261 | G | A | 0.371 | 0.039 | 1.04 | 4.83E-08 | Bellenguez | Microglia | 0.153959842 |
| rs7401792 | SLC24A4 | 14 | 92931261 | G | A | 0.371 | 0.039 | 1.04 | 4.83E-08 | Bellenguez | Oligodendrocytes | 0.001439572 |
| rs7401792 | SLC24A4 | 14 | 92931261 | G | A | 0.371 | 0.039 | 1.04 | 4.83E-08 | Bellenguez | OPC | 0.066999633 |
| rs12590654 | SLC24A4 | 14 | 92938855 | G | A | 0.672 | 0.062 | 1.06 | 4.25E-21 | Bellenguez | Astrocytes | 0.156636464 |
| rs12590654 | SLC24A4 | 14 | 92938855 | G | A | 0.672 | 0.062 | 1.06 | 4.25E-21 | Bellenguez | Endothelial | 0 |
| rs12590654 | SLC24A4 | 14 | 92938855 | G | A | 0.672 | 0.062 | 1.06 | 4.25E-21 | Bellenguez | GABAergic | 0.337108325 |
| rs12590654 | SLC24A4 | 14 | 92938855 | G | A | 0.672 | 0.062 | 1.06 | 4.25E-21 | Bellenguez | Glutamatergic | 0.283856164 |
| rs12590654 | SLC24A4 | 14 | 92938855 | G | A | 0.672 | 0.062 | 1.06 | 4.25E-21 | Bellenguez | Microglia | 0.153959842 |
| rs12590654 | SLC24A4 | 14 | 92938855 | G | A | 0.672 | 0.062 | 1.06 | 4.25E-21 | Bellenguez | Oligodendrocytes | 0.001439572 |
| rs12590654 | SLC24A4 | 14 | 92938855 | G | A | 0.672 | 0.062 | 1.06 | 4.25E-21 | Bellenguez | OPC | 0.066999633 |
| rs8025980 | SPPL2A | 15 | 50994011 | A | G | 0.655 | 0.062 | 1.06 | 1.32E-08 | Bellenguez | Astrocytes | 0.099010608 |
| rs8025980 | SPPL2A | 15 | 50994011 | A | G | 0.655 | 0.062 | 1.06 | 1.32E-08 | Bellenguez | Endothelial | 0.057941107 |

| | | | | | | | | | | | | |
|---|---|---|---|---|---|---|---|---|---|---|---|---|
| rs8025980 | SPPL2A | 15 | 50994011 | A | G | 0.655 | 0.062 | 1.06 | 1.32E-08 | Bellenguez | GABAergic | 0.039200942 |
| rs8025980 | SPPL2A | 15 | 50994011 | A | G | 0.655 | 0.062 | 1.06 | 1.32E-08 | Bellenguez | Glutamatergic | 0.038865702 |
| rs8025980 | SPPL2A | 15 | 50994011 | A | G | 0.655 | 0.062 | 1.06 | 1.32E-08 | Bellenguez | Microglia | 0.685367241 |
| rs8025980 | SPPL2A | 15 | 50994011 | A | G | 0.655 | 0.062 | 1.06 | 1.32E-08 | Bellenguez | Oligodendrocytes | 0.026444326 |
| rs8025980 | SPPL2A | 15 | 50994011 | A | G | 0.655 | 0.062 | 1.06 | 1.32E-08 | Bellenguez | OPC | 0.053170074 |
| rs117618017 | APH1B | 15 | 63569902 | T | C | 0.144 | 0.086 | 1.09 | 2.15E-25 | Bellenguez | Astrocytes | 0.132311333 |
| rs117618017 | APH1B | 15 | 63569902 | T | C | 0.144 | 0.086 | 1.09 | 2.15E-25 | Bellenguez | Endothelial | 0 |
| rs117618017 | APH1B | 15 | 63569902 | T | C | 0.144 | 0.086 | 1.09 | 2.15E-25 | Bellenguez | GABAergic | 0.140578299 |
| rs117618017 | APH1B | 15 | 63569902 | T | C | 0.144 | 0.086 | 1.09 | 2.15E-25 | Bellenguez | Glutamatergic | 0.081955756 |
| rs117618017 | APH1B | 15 | 63569902 | T | C | 0.144 | 0.086 | 1.09 | 2.15E-25 | Bellenguez | Microglia | 0.333812466 |
| rs117618017 | APH1B | 15 | 63569902 | T | C | 0.144 | 0.086 | 1.09 | 2.15E-25 | Bellenguez | Oligodendrocytes | 0.083442752 |
| rs117618017 | APH1B | 15 | 63569902 | T | C | 0.144 | 0.086 | 1.09 | 2.15E-25 | Bellenguez | OPC | 0.227899395 |
| rs3848143 | SNX1 | 15 | 64423506 | G | A | 0.22 | 0.068 | 1.07 | 8.41E-11 | Bellenguez | Astrocytes | 0.067168853 |
| rs3848143 | SNX1 | 15 | 64423506 | G | A | 0.22 | 0.068 | 1.07 | 8.41E-11 | Bellenguez | Endothelial | 0.000615136 |
| rs3848143 | SNX1 | 15 | 64423506 | G | A | 0.22 | 0.068 | 1.07 | 8.41E-11 | Bellenguez | GABAergic | 0.077002393 |
| rs3848143 | SNX1 | 15 | 64423506 | G | A | 0.22 | 0.068 | 1.07 | 8.41E-11 | Bellenguez | Glutamatergic | 0.074435513 |
| rs3848143 | SNX1 | 15 | 64423506 | G | A | 0.22 | 0.068 | 1.07 | 8.41E-11 | Bellenguez | Microglia | 0.132264935 |
| rs3848143 | SNX1 | 15 | 64423506 | G | A | 0.22 | 0.068 | 1.07 | 8.41E-11 | Bellenguez | Oligodendrocytes | 0.357576915 |
| rs3848143 | SNX1 | 15 | 64423506 | G | A | 0.22 | 0.068 | 1.07 | 8.41E-11 | Bellenguez | OPC | 0.290936255 |
| rs12592898 | CTSH | 15 | 79229199 | G | A | 0.867 | 0.062 | 1.06 | 4.18E-09 | Bellenguez | Astrocytes | 0.499425454 |
| rs12592898 | CTSH | 15 | 79229199 | G | A | 0.867 | 0.062 | 1.06 | 4.18E-09 | Bellenguez | Endothelial | 0.006189521 |
| rs12592898 | CTSH | 15 | 79229199 | G | A | 0.867 | 0.062 | 1.06 | 4.18E-09 | Bellenguez | GABAergic | 0.031427726 |
| rs12592898 | CTSH | 15 | 79229199 | G | A | 0.867 | 0.062 | 1.06 | 4.18E-09 | Bellenguez | Glutamatergic | 0.018462082 |
| rs12592898 | CTSH | 15 | 79229199 | G | A | 0.867 | 0.062 | 1.06 | 4.18E-09 | Bellenguez | Microglia | 0.434621975 |
| rs12592898 | CTSH | 15 | 79229199 | G | A | 0.867 | 0.062 | 1.06 | 4.18E-09 | Bellenguez | Oligodendrocytes | 0.00075457 |
| rs12592898 | CTSH | 15 | 79229199 | G | A | 0.867 | 0.062 | 1.06 | 4.18E-09 | Bellenguez | OPC | 0.009118672 |
| rs889555 | BCKDK | 16 | 31122571 | C | T | 0.719 | 0.041 | 1.04 | 1.96E-11 | Bellenguez | Astrocytes | 0.224050883 |
| rs889555 | BCKDK | 16 | 31122571 | C | T | 0.719 | 0.041 | 1.04 | 1.96E-11 | Bellenguez | Endothelial | 0.221819216 |
| rs889555 | BCKDK | 16 | 31122571 | C | T | 0.719 | 0.041 | 1.04 | 1.96E-11 | Bellenguez | GABAergic | 0.125863192 |
| rs889555 | BCKDK | 16 | 31122571 | C | T | 0.719 | 0.041 | 1.04 | 1.96E-11 | Bellenguez | Glutamatergic | 0.117758585 |
| rs889555 | BCKDK | 16 | 31122571 | C | T | 0.719 | 0.041 | 1.04 | 1.96E-11 | Bellenguez | Microglia | 0.00687632 |
| rs889555 | BCKDK | 16 | 31122571 | C | T | 0.719 | 0.041 | 1.04 | 1.96E-11 | Bellenguez | Oligodendrocytes | 0.142769413 |
| rs889555 | BCKDK | 16 | 31122571 | C | T | 0.719 | 0.041 | 1.04 | 1.96E-11 | Bellenguez | OPC | 0.16086239 |
| rs4985556 | IL34 | 16 | 70694000 | A | C | 0.115 | 0.086 | 1.09 | 5.98E-10 | Bellenguez | Astrocytes | 0.005640773 |
| rs4985556 | IL34 | 16 | 70694000 | A | C | 0.115 | 0.086 | 1.09 | 5.98E-10 | Bellenguez | Endothelial | 0 |

| | | | | | | | | | | | | |
|---|---|---|---|---|---|---|---|---|---|---|---|---|
| rs4985556 | IL34 | 16 | 70694000 | A | C | 0.115 | 0.086 | 1.09 | 5.98E-10 | Bellenguez | GABAergic | 0.43062314 |
| rs4985556 | IL34 | 16 | 70694000 | A | C | 0.115 | 0.086 | 1.09 | 5.98E-10 | Bellenguez | Glutamatergic | 0.524891129 |
| rs4985556 | IL34 | 16 | 70694000 | A | C | 0.115 | 0.086 | 1.09 | 5.98E-10 | Bellenguez | Microglia | 0.036619678 |
| rs4985556 | IL34 | 16 | 70694000 | A | C | 0.115 | 0.086 | 1.09 | 5.98E-10 | Bellenguez | Oligodendrocytes | 0.000731986 |
| rs4985556 | IL34 | 16 | 70694000 | A | C | 0.115 | 0.086 | 1.09 | 5.98E-10 | Bellenguez | OPC | 0.001493294 |
| rs450674 | MAF | 16 | 79608408 | T | C | 0.627 | 0.020 | 1.02 | 3.16E-08 | Bellenguez | Astrocytes | 0.041136125 |
| rs450674 | MAF | 16 | 79608408 | T | C | 0.627 | 0.020 | 1.02 | 3.16E-08 | Bellenguez | Endothelial | 0.023971481 |
| rs450674 | MAF | 16 | 79608408 | T | C | 0.627 | 0.020 | 1.02 | 3.16E-08 | Bellenguez | GABAergic | 0.093989451 |
| rs450674 | MAF | 16 | 79608408 | T | C | 0.627 | 0.020 | 1.02 | 3.16E-08 | Bellenguez | Glutamatergic | 0.001008411 |
| rs450674 | MAF | 16 | 79608408 | T | C | 0.627 | 0.020 | 1.02 | 3.16E-08 | Bellenguez | Microglia | 0.748275058 |
| rs450674 | MAF | 16 | 79608408 | T | C | 0.627 | 0.020 | 1.02 | 3.16E-08 | Bellenguez | Oligodendrocytes | 0.022982328 |
| rs450674 | MAF | 16 | 79608408 | T | C | 0.627 | 0.020 | 1.02 | 3.16E-08 | Bellenguez | OPC | 0.068637146 |
| rs12446759 | PLCG2 | 16 | 81773003 | A | G | 0.597 | 0.041 | 1.04 | 1.22E-13 | Bellenguez | Astrocytes | 0.005300271 |
| rs12446759 | PLCG2 | 16 | 81773003 | A | G | 0.597 | 0.041 | 1.04 | 1.22E-13 | Bellenguez | Endothelial | 0.15557428 |
| rs12446759 | PLCG2 | 16 | 81773003 | A | G | 0.597 | 0.041 | 1.04 | 1.22E-13 | Bellenguez | GABAergic | 0.00427693 |
| rs12446759 | PLCG2 | 16 | 81773003 | A | G | 0.597 | 0.041 | 1.04 | 1.22E-13 | Bellenguez | Glutamatergic | 0.001800433 |
| rs12446759 | PLCG2 | 16 | 81773003 | A | G | 0.597 | 0.041 | 1.04 | 1.22E-13 | Bellenguez | Microglia | 0.748216032 |
| rs12446759 | PLCG2 | 16 | 81773003 | A | G | 0.597 | 0.041 | 1.04 | 1.22E-13 | Bellenguez | Oligodendrocytes | 0.021528744 |
| rs12446759 | PLCG2 | 16 | 81773003 | A | G | 0.597 | 0.041 | 1.04 | 1.22E-13 | Bellenguez | OPC | 0.06330331 |
| rs72824905 | PLCG2 | 16 | 81942028 | C | G | 0.992 | 0.248 | 1.28 | 8.48E-12 | Bellenguez | Astrocytes | 0.005300271 |
| rs72824905 | PLCG2 | 16 | 81942028 | C | G | 0.992 | 0.248 | 1.28 | 8.48E-12 | Bellenguez | Endothelial | 0.15557428 |
| rs72824905 | PLCG2 | 16 | 81942028 | C | G | 0.992 | 0.248 | 1.28 | 8.48E-12 | Bellenguez | GABAergic | 0.00427693 |
| rs72824905 | PLCG2 | 16 | 81942028 | C | G | 0.992 | 0.248 | 1.28 | 8.48E-12 | Bellenguez | Glutamatergic | 0.001800433 |
| rs72824905 | PLCG2 | 16 | 81942028 | C | G | 0.992 | 0.248 | 1.28 | 8.48E-12 | Bellenguez | Microglia | 0.748216032 |
| rs72824905 | PLCG2 | 16 | 81942028 | C | G | 0.992 | 0.248 | 1.28 | 8.48E-12 | Bellenguez | Oligodendrocytes | 0.021528744 |
| rs72824905 | PLCG2 | 16 | 81942028 | C | G | 0.992 | 0.248 | 1.28 | 8.48E-12 | Bellenguez | OPC | 0.06330331 |
| rs16941239 | FOXF1 | 16 | 86454210 | A | T | 0.029 | 0.113 | 1.12 | 1.29E-08 | Bellenguez | Astrocytes | 4.10094E-05 |
| rs16941239 | FOXF1 | 16 | 86454210 | A | T | 0.029 | 0.113 | 1.12 | 1.29E-08 | Bellenguez | Endothelial | 0.999286111 |
| rs16941239 | FOXF1 | 16 | 86454210 | A | T | 0.029 | 0.113 | 1.12 | 1.29E-08 | Bellenguez | GABAergic | 0.000433351 |
| rs16941239 | FOXF1 | 16 | 86454210 | A | T | 0.029 | 0.113 | 1.12 | 1.29E-08 | Bellenguez | Glutamatergic | 0.000239529 |
| rs16941239 | FOXF1 | 16 | 86454210 | A | T | 0.029 | 0.113 | 1.12 | 1.29E-08 | Bellenguez | Microglia | 0 |
| rs16941239 | FOXF1 | 16 | 86454210 | A | T | 0.029 | 0.113 | 1.12 | 1.29E-08 | Bellenguez | Oligodendrocytes | 0 |
| rs16941239 | FOXF1 | 16 | 86454210 | A | T | 0.029 | 0.113 | 1.12 | 1.29E-08 | Bellenguez | OPC | 0 |
| rs56407236 | PRDM7 | 16 | 90170095 | A | G | 0.069 | 0.104 | 1.11 | 6.47E-15 | Bellenguez | Astrocytes | 0 |
| rs56407236 | PRDM7 | 16 | 90170095 | A | G | 0.069 | 0.104 | 1.11 | 6.47E-15 | Bellenguez | Endothelial | 0 |

| | | | | | | | | | | | | |
|---|---|---|---|---|---|---|---|---|---|---|---|---|
| rs56407236 | PRDM7 | 16 | 90170095 | A | G | 0.069 | 0.104 | 1.11 | 6.47E-15 | Bellenguez | GABAergic | 0.424139996 |
| rs56407236 | PRDM7 | 16 | 90170095 | A | G | 0.069 | 0.104 | 1.11 | 6.47E-15 | Bellenguez | Glutamatergic | 0.342223504 |
| rs56407236 | PRDM7 | 16 | 90170095 | A | G | 0.069 | 0.104 | 1.11 | 6.47E-15 | Bellenguez | Microglia | 0 |
| rs56407236 | PRDM7 | 16 | 90170095 | A | G | 0.069 | 0.104 | 1.11 | 6.47E-15 | Bellenguez | Oligodendrocytes | 0.2336365 |
| rs56407236 | PRDM7 | 16 | 90170095 | A | G | 0.069 | 0.104 | 1.11 | 6.47E-15 | Bellenguez | OPC | 0 |
| rs2242595 | MYO15A | 17 | 18059454 | G | A | 0.888 | 0.073 | 1.08 | 1.11E-09 | Bellenguez | Astrocytes | 0.092481018 |
| rs2242595 | MYO15A | 17 | 18059454 | G | A | 0.888 | 0.073 | 1.08 | 1.11E-09 | Bellenguez | Endothelial | 0 |
| rs2242595 | MYO15A | 17 | 18059454 | G | A | 0.888 | 0.073 | 1.08 | 1.11E-09 | Bellenguez | GABAergic | 0.285503634 |
| rs2242595 | MYO15A | 17 | 18059454 | G | A | 0.888 | 0.073 | 1.08 | 1.11E-09 | Bellenguez | Glutamatergic | 0.34259131 |
| rs2242595 | MYO15A | 17 | 18059454 | G | A | 0.888 | 0.073 | 1.08 | 1.11E-09 | Bellenguez | Microglia | 0.027224007 |
| rs2242595 | MYO15A | 17 | 18059454 | G | A | 0.888 | 0.073 | 1.08 | 1.11E-09 | Bellenguez | Oligodendrocytes | 0.2515853 |
| rs2242595 | MYO15A | 17 | 18059454 | G | A | 0.888 | 0.073 | 1.08 | 1.11E-09 | Bellenguez | OPC | 0.000614731 |
| rs5848 | GRN | 17 | 42430244 | T | C | 0.289 | 0.077 | 1.08 | 2.38E-20 | Bellenguez | Astrocytes | 0.041895715 |
| rs5848 | GRN | 17 | 42430244 | T | C | 0.289 | 0.077 | 1.08 | 2.38E-20 | Bellenguez | Endothelial | 0.086565116 |
| rs5848 | GRN | 17 | 42430244 | T | C | 0.289 | 0.077 | 1.08 | 2.38E-20 | Bellenguez | GABAergic | 0.048265534 |
| rs5848 | GRN | 17 | 42430244 | T | C | 0.289 | 0.077 | 1.08 | 2.38E-20 | Bellenguez | Glutamatergic | 0.032869867 |
| rs5848 | GRN | 17 | 42430244 | T | C | 0.289 | 0.077 | 1.08 | 2.38E-20 | Bellenguez | Microglia | 0.616795386 |
| rs5848 | GRN | 17 | 42430244 | T | C | 0.289 | 0.077 | 1.08 | 2.38E-20 | Bellenguez | Oligodendrocytes | 0.100953438 |
| rs5848 | GRN | 17 | 42430244 | T | C | 0.289 | 0.077 | 1.08 | 2.38E-20 | Bellenguez | OPC | 0.072654943 |
| rs199515 | WNT3 | 17 | 44856641 | C | G | 0.781 | 0.073 | 1.08 | 9.34E-13 | Bellenguez | Astrocytes | 0.041406074 |
| rs199515 | WNT3 | 17 | 44856641 | C | G | 0.781 | 0.073 | 1.08 | 9.34E-13 | Bellenguez | Endothelial | 0 |
| rs199515 | WNT3 | 17 | 44856641 | C | G | 0.781 | 0.073 | 1.08 | 9.34E-13 | Bellenguez | GABAergic | 0.400162619 |
| rs199515 | WNT3 | 17 | 44856641 | C | G | 0.781 | 0.073 | 1.08 | 9.34E-13 | Bellenguez | Glutamatergic | 0.303226393 |
| rs199515 | WNT3 | 17 | 44856641 | C | G | 0.781 | 0.073 | 1.08 | 9.34E-13 | Bellenguez | Microglia | 0 |
| rs199515 | WNT3 | 17 | 44856641 | C | G | 0.781 | 0.073 | 1.08 | 9.34E-13 | Bellenguez | Oligodendrocytes | 0.006049521 |
| rs199515 | WNT3 | 17 | 44856641 | C | G | 0.781 | 0.073 | 1.08 | 9.34E-13 | Bellenguez | OPC | 0.249155394 |
| rs28394864 | ZNF652 | 17 | 47450775 | A | G | 0.46 | 0.009 | 1.01 | 4.9E-10 | Wightman | Astrocytes | 0.081139067 |
| rs28394864 | ZNF652 | 17 | 47450775 | A | G | 0.46 | 0.009 | 1.01 | 4.9E-10 | Wightman | Endothelial | 0.082850358 |
| rs28394864 | ZNF652 | 17 | 47450775 | A | G | 0.46 | 0.009 | 1.01 | 4.9E-10 | Wightman | GABAergic | 0.096814082 |
| rs28394864 | ZNF652 | 17 | 47450775 | A | G | 0.46 | 0.009 | 1.01 | 4.9E-10 | Wightman | Glutamatergic | 0.063536168 |
| rs28394864 | ZNF652 | 17 | 47450775 | A | G | 0.46 | 0.009 | 1.01 | 4.9E-10 | Wightman | Microglia | 0.271905122 |
| rs28394864 | ZNF652 | 17 | 47450775 | A | G | 0.46 | 0.009 | 1.01 | 4.9E-10 | Wightman | Oligodendrocytes | 0.270235922 |
| rs28394864 | ZNF652 | 17 | 47450775 | A | G | 0.46 | 0.009 | 1.01 | 4.9E-10 | Wightman | OPC | 0.133519281 |
| rs7225151 | SCIMP | 17 | 5137047 | A | G | 0.124 | 0.049 | 1.05 | 4.13E-13 | Bellenguez | Astrocytes | 0.026077185 |
| rs7225151 | SCIMP | 17 | 5137047 | A | G | 0.124 | 0.049 | 1.05 | 4.13E-13 | Bellenguez | Endothelial | 0 |

| | | | | | | | | | | | | |
|---|---|---|---|---|---|---|---|---|---|---|---|---|
| rs7225151 | SCIMP | 17 | 5137047 | A | G | 0.124 | 0.049 | 1.05 | 4.13E-13 | Bellenguez | GABAergic | 0.095043897 |
| rs7225151 | SCIMP | 17 | 5137047 | A | G | 0.124 | 0.049 | 1.05 | 4.13E-13 | Bellenguez | Glutamatergic | 0.11846687 |
| rs7225151 | SCIMP | 17 | 5137047 | A | G | 0.124 | 0.049 | 1.05 | 4.13E-13 | Bellenguez | Microglia | 0.678828879 |
| rs7225151 | SCIMP | 17 | 5137047 | A | G | 0.124 | 0.049 | 1.05 | 4.13E-13 | Bellenguez | Oligodendrocytes | 0.048060513 |
| rs7225151 | SCIMP | 17 | 5137047 | A | G | 0.124 | 0.049 | 1.05 | 4.13E-13 | Bellenguez | OPC | 0.033522656 |
| rs4277405 | ACE | 17 | 61548918 | T | C | 0.616 | 0.051 | 1.05 | 8.8E-20 | Bellenguez | Astrocytes | 0.033462093 |
| rs4277405 | ACE | 17 | 61548918 | T | C | 0.616 | 0.051 | 1.05 | 8.8E-20 | Bellenguez | Endothelial | 0 |
| rs4277405 | ACE | 17 | 61548918 | T | C | 0.616 | 0.051 | 1.05 | 8.8E-20 | Bellenguez | GABAergic | 0.334887256 |
| rs4277405 | ACE | 17 | 61548918 | T | C | 0.616 | 0.051 | 1.05 | 8.8E-20 | Bellenguez | Glutamatergic | 0.610248373 |
| rs4277405 | ACE | 17 | 61548918 | T | C | 0.616 | 0.051 | 1.05 | 8.8E-20 | Bellenguez | Microglia | 0 |
| rs4277405 | ACE | 17 | 61548918 | T | C | 0.616 | 0.051 | 1.05 | 8.8E-20 | Bellenguez | Oligodendrocytes | 0.002115702 |
| rs4277405 | ACE | 17 | 61548918 | T | C | 0.616 | 0.051 | 1.05 | 8.8E-20 | Bellenguez | OPC | 0.019286575 |
| rs12151021 | ABCA7 | 19 | 1050874 | A | G | 0.336 | 0.086 | 1.09 | 1.59E-37 | Bellenguez | Astrocytes | 0.175805931 |
| rs12151021 | ABCA7 | 19 | 1050874 | A | G | 0.336 | 0.086 | 1.09 | 1.59E-37 | Bellenguez | Endothelial | 0.204717928 |
| rs12151021 | ABCA7 | 19 | 1050874 | A | G | 0.336 | 0.086 | 1.09 | 1.59E-37 | Bellenguez | GABAergic | 0.210055663 |
| rs12151021 | ABCA7 | 19 | 1050874 | A | G | 0.336 | 0.086 | 1.09 | 1.59E-37 | Bellenguez | Glutamatergic | 0.243133589 |
| rs12151021 | ABCA7 | 19 | 1050874 | A | G | 0.336 | 0.086 | 1.09 | 1.59E-37 | Bellenguez | Microglia | 0.144390338 |
| rs12151021 | ABCA7 | 19 | 1050874 | A | G | 0.336 | 0.086 | 1.09 | 1.59E-37 | Bellenguez | Oligodendrocytes | 0.021851088 |
| rs12151021 | ABCA7 | 19 | 1050874 | A | G | 0.336 | 0.086 | 1.09 | 1.59E-37 | Bellenguez | OPC | 4.54618E-05 |
| rs2452170 | MAMSTR | 19 | 49213504 | A | G | 0.53 | 0.008 | 1.01 | 1.72E-08 | Wightman | Astrocytes | 0.346461906 |
| rs2452170 | MAMSTR | 19 | 49213504 | A | G | 0.53 | 0.008 | 1.01 | 1.72E-08 | Wightman | Endothelial | 0 |
| rs2452170 | MAMSTR | 19 | 49213504 | A | G | 0.53 | 0.008 | 1.01 | 1.72E-08 | Wightman | GABAergic | 0.172929059 |
| rs2452170 | MAMSTR | 19 | 49213504 | A | G | 0.53 | 0.008 | 1.01 | 1.72E-08 | Wightman | Glutamatergic | 0.232339861 |
| rs2452170 | MAMSTR | 19 | 49213504 | A | G | 0.53 | 0.008 | 1.01 | 1.72E-08 | Wightman | Microglia | 0 |
| rs2452170 | MAMSTR | 19 | 49213504 | A | G | 0.53 | 0.008 | 1.01 | 1.72E-08 | Wightman | Oligodendrocytes | 0.003740308 |
| rs2452170 | MAMSTR | 19 | 49213504 | A | G | 0.53 | 0.008 | 1.01 | 1.72E-08 | Wightman | OPC | 0.244528865 |
| rs9304690 | SIGLEC11 | 19 | 50453317 | T | C | 0.24 | 0.058 | 1.06 | 4.74E-09 | Bellenguez | Astrocytes | 0.011977987 |
| rs9304690 | SIGLEC11 | 19 | 50453317 | T | C | 0.24 | 0.058 | 1.06 | 4.74E-09 | Bellenguez | Endothelial | 0 |
| rs9304690 | SIGLEC11 | 19 | 50453317 | T | C | 0.24 | 0.058 | 1.06 | 4.74E-09 | Bellenguez | GABAergic | 0.014620799 |
| rs9304690 | SIGLEC11 | 19 | 50453317 | T | C | 0.24 | 0.058 | 1.06 | 4.74E-09 | Bellenguez | Glutamatergic | 0.013612762 |
| rs9304690 | SIGLEC11 | 19 | 50453317 | T | C | 0.24 | 0.058 | 1.06 | 4.74E-09 | Bellenguez | Microglia | 0.945618323 |
| rs9304690 | SIGLEC11 | 19 | 50453317 | T | C | 0.24 | 0.058 | 1.06 | 4.74E-09 | Bellenguez | Oligodendrocytes | 0.010170934 |
| rs9304690 | SIGLEC11 | 19 | 50453317 | T | C | 0.24 | 0.058 | 1.06 | 4.74E-09 | Bellenguez | OPC | 0.003999195 |
| rs1354106 | CD33 | 19 | 51737991 | T | G | 0.63 | 0.011 | 1.01 | 2.21E-10 | Wightman | Astrocytes | 0.004493026 |
| rs1354106 | CD33 | 19 | 51737991 | T | G | 0.63 | 0.011 | 1.01 | 2.21E-10 | Wightman | Endothelial | 0 |

| | | | | | | | | | | | | |
|---|---|---|---|---|---|---|---|---|---|---|---|---|
| rs1354106 | CD33 | 19 | 51737991 | T | G | 0.63 | 0.011 | 1.01 | 2.21E-10 | Wightman | GABAergic | 0.00077343 |
| rs1354106 | CD33 | 19 | 51737991 | T | G | 0.63 | 0.011 | 1.01 | 2.21E-10 | Wightman | Glutamatergic | 0.000877081 |
| rs1354106 | CD33 | 19 | 51737991 | T | G | 0.63 | 0.011 | 1.01 | 2.21E-10 | Wightman | Microglia | 0.935109646 |
| rs1354106 | CD33 | 19 | 51737991 | T | G | 0.63 | 0.011 | 1.01 | 2.21E-10 | Wightman | Oligodendrocytes | 0.058638661 |
| rs1354106 | CD33 | 19 | 51737991 | T | G | 0.63 | 0.011 | 1.01 | 2.21E-10 | Wightman | OPC | 0.000108157 |
| rs587709 | LILRB2 | 19 | 54771451 | C | T | 0.325 | 0.049 | 1.05 | 3.63E-11 | Bellenguez | Astrocytes | 0.000807069 |
| rs587709 | LILRB2 | 19 | 54771451 | C | T | 0.325 | 0.049 | 1.05 | 3.63E-11 | Bellenguez | Endothelial | 0 |
| rs587709 | LILRB2 | 19 | 54771451 | C | T | 0.325 | 0.049 | 1.05 | 3.63E-11 | Bellenguez | GABAergic | 0.00077824 |
| rs587709 | LILRB2 | 19 | 54771451 | C | T | 0.325 | 0.049 | 1.05 | 3.63E-11 | Bellenguez | Glutamatergic | 0.000842904 |
| rs587709 | LILRB2 | 19 | 54771451 | C | T | 0.325 | 0.049 | 1.05 | 3.63E-11 | Bellenguez | Microglia | 0.996795082 |
| rs587709 | LILRB2 | 19 | 54771451 | C | T | 0.325 | 0.049 | 1.05 | 3.63E-11 | Bellenguez | Oligodendrocytes | 0 |
| rs587709 | LILRB2 | 19 | 54771451 | C | T | 0.325 | 0.049 | 1.05 | 3.63E-11 | Bellenguez | OPC | 0.000776704 |
| rs1761461 | LILRA5 | 19 | 54825174 | C | A | 0.49 | 0.008 | 1.01 | 1.56E-09 | Wightman | Astrocytes | 0.131957271 |
| rs1761461 | LILRA5 | 19 | 54825174 | C | A | 0.49 | 0.008 | 1.01 | 1.56E-09 | Wightman | Endothelial | 0 |
| rs1761461 | LILRA5 | 19 | 54825174 | C | A | 0.49 | 0.008 | 1.01 | 1.56E-09 | Wightman | GABAergic | 0.277340611 |
| rs1761461 | LILRA5 | 19 | 54825174 | C | A | 0.49 | 0.008 | 1.01 | 1.56E-09 | Wightman | Glutamatergic | 0.24636203 |
| rs1761461 | LILRA5 | 19 | 54825174 | C | A | 0.49 | 0.008 | 1.01 | 1.56E-09 | Wightman | Microglia | 0.183087584 |
| rs1761461 | LILRA5 | 19 | 54825174 | C | A | 0.49 | 0.008 | 1.01 | 1.56E-09 | Wightman | Oligodendrocytes | 0.082271867 |
| rs1761461 | LILRA5 | 19 | 54825174 | C | A | 0.49 | 0.008 | 1.01 | 1.56E-09 | Wightman | OPC | 0.078980638 |
| rs141749679 | SORT1 | 1 | 109888432 | C | T | 0.004 | 0.344 | 1.41 | 7.54E-09 | Bellenguez | Astrocytes | 0.055539985 |
| rs141749679 | SORT1 | 1 | 109888432 | C | T | 0.004 | 0.344 | 1.41 | 7.54E-09 | Bellenguez | Endothelial | 0.133177665 |
| rs141749679 | SORT1 | 1 | 109888432 | C | T | 0.004 | 0.344 | 1.41 | 7.54E-09 | Bellenguez | GABAergic | 0.068982486 |
| rs141749679 | SORT1 | 1 | 109888432 | C | T | 0.004 | 0.344 | 1.41 | 7.54E-09 | Bellenguez | Glutamatergic | 0.069593001 |
| rs141749679 | SORT1 | 1 | 109888432 | C | T | 0.004 | 0.344 | 1.41 | 7.54E-09 | Bellenguez | Microglia | 0.091652498 |
| rs141749679 | SORT1 | 1 | 109888432 | C | T | 0.004 | 0.344 | 1.41 | 7.54E-09 | Bellenguez | Oligodendrocytes | 0.510537393 |
| rs141749679 | SORT1 | 1 | 109888432 | C | T | 0.004 | 0.344 | 1.41 | 7.54E-09 | Bellenguez | OPC | 0.070516973 |
| rs679515 | CR1 | 1 | 207750568 | T | C | 0.188 | 0.113 | 1.12 | 7.16E-46 | Bellenguez | Astrocytes | 0.00204073 |
| rs679515 | CR1 | 1 | 207750568 | T | C | 0.188 | 0.113 | 1.12 | 7.16E-46 | Bellenguez | Endothelial | 0 |
| rs679515 | CR1 | 1 | 207750568 | T | C | 0.188 | 0.113 | 1.12 | 7.16E-46 | Bellenguez | GABAergic | 0.019285076 |
| rs679515 | CR1 | 1 | 207750568 | T | C | 0.188 | 0.113 | 1.12 | 7.16E-46 | Bellenguez | Glutamatergic | 0.019542727 |
| rs679515 | CR1 | 1 | 207750568 | T | C | 0.188 | 0.113 | 1.12 | 7.16E-46 | Bellenguez | Microglia | 0 |
| rs679515 | CR1 | 1 | 207750568 | T | C | 0.188 | 0.113 | 1.12 | 7.16E-46 | Bellenguez | Oligodendrocytes | 0.959131466 |
| rs679515 | CR1 | 1 | 207750568 | T | C | 0.188 | 0.113 | 1.12 | 7.16E-46 | Bellenguez | OPC | 0 |
| rs113020870 | AGRN | 1 | 985377 | T | C | 0.0041 | 0.069 | 1.07 | 3.83E-08 | Wightman | Astrocytes | 0.184020054 |
| rs113020870 | AGRN | 1 | 985377 | T | C | 0.0041 | 0.069 | 1.07 | 3.83E-08 | Wightman | Endothelial | 0.021139675 |

| | | | | | | | | | | | | |
|---|---|---|---|---|---|---|---|---|---|---|---|---|
| rs113020870 | AGRN | 1 | 985377 | T | C | 0.0041 | 0.069 | 1.07 | 3.83E-08 | Wightman | GABAergic | 0.077826931 |
| rs113020870 | AGRN | 1 | 985377 | T | C | 0.0041 | 0.069 | 1.07 | 3.83E-08 | Wightman | Glutamatergic | 0.182818072 |
| rs113020870 | AGRN | 1 | 985377 | T | C | 0.0041 | 0.069 | 1.07 | 3.83E-08 | Wightman | Microglia | 0.003480236 |
| rs113020870 | AGRN | 1 | 985377 | T | C | 0.0041 | 0.069 | 1.07 | 3.83E-08 | Wightman | Oligodendrocytes | 0.328020268 |
| rs113020870 | AGRN | 1 | 985377 | T | C | 0.0041 | 0.069 | 1.07 | 3.83E-08 | Wightman | OPC | 0.202694764 |
| rs1358782 | RBCK1 | 20 | 393978 | G | A | 0.754 | 0.041 | 1.04 | 1.55E-08 | Bellenguez | Astrocytes | 0.243041267 |
| rs1358782 | RBCK1 | 20 | 393978 | G | A | 0.754 | 0.041 | 1.04 | 1.55E-08 | Bellenguez | Endothelial | 0.154651983 |
| rs1358782 | RBCK1 | 20 | 393978 | G | A | 0.754 | 0.041 | 1.04 | 1.55E-08 | Bellenguez | GABAergic | 0.096350059 |
| rs1358782 | RBCK1 | 20 | 393978 | G | A | 0.754 | 0.041 | 1.04 | 1.55E-08 | Bellenguez | Glutamatergic | 0.070456883 |
| rs1358782 | RBCK1 | 20 | 393978 | G | A | 0.754 | 0.041 | 1.04 | 1.55E-08 | Bellenguez | Microglia | 0.166344658 |
| rs1358782 | RBCK1 | 20 | 393978 | G | A | 0.754 | 0.041 | 1.04 | 1.55E-08 | Bellenguez | Oligodendrocytes | 0.090951466 |
| rs1358782 | RBCK1 | 20 | 393978 | G | A | 0.754 | 0.041 | 1.04 | 1.55E-08 | Bellenguez | OPC | 0.178203682 |
| rs6014724 | CASS4 | 20 | 54998544 | A | G | 0.91 | 0.117 | 1.12 | 4.13E-21 | Bellenguez | Astrocytes | 0.000429865 |
| rs6014724 | CASS4 | 20 | 54998544 | A | G | 0.91 | 0.117 | 1.12 | 4.13E-21 | Bellenguez | Endothelial | 0.636316788 |
| rs6014724 | CASS4 | 20 | 54998544 | A | G | 0.91 | 0.117 | 1.12 | 4.13E-21 | Bellenguez | GABAergic | 0.00167726 |
| rs6014724 | CASS4 | 20 | 54998544 | A | G | 0.91 | 0.117 | 1.12 | 4.13E-21 | Bellenguez | Glutamatergic | 0.002014677 |
| rs6014724 | CASS4 | 20 | 54998544 | A | G | 0.91 | 0.117 | 1.12 | 4.13E-21 | Bellenguez | Microglia | 0.357060287 |
| rs6014724 | CASS4 | 20 | 54998544 | A | G | 0.91 | 0.117 | 1.12 | 4.13E-21 | Bellenguez | Oligodendrocytes | 0.002207992 |
| rs6014724 | CASS4 | 20 | 54998544 | A | G | 0.91 | 0.117 | 1.12 | 4.13E-21 | Bellenguez | OPC | 0.000293131 |
| rs6742 | SLC2A4RG | 20 | 62374441 | C | T | 0.779 | 0.062 | 1.06 | 2.58E-09 | Bellenguez | Astrocytes | 0.433262639 |
| rs6742 | SLC2A4RG | 20 | 62374441 | C | T | 0.779 | 0.062 | 1.06 | 2.58E-09 | Bellenguez | Endothelial | 0.056237308 |
| rs6742 | SLC2A4RG | 20 | 62374441 | C | T | 0.779 | 0.062 | 1.06 | 2.58E-09 | Bellenguez | GABAergic | 0.174500789 |
| rs6742 | SLC2A4RG | 20 | 62374441 | C | T | 0.779 | 0.062 | 1.06 | 2.58E-09 | Bellenguez | Glutamatergic | 0.103253958 |
| rs6742 | SLC2A4RG | 20 | 62374441 | C | T | 0.779 | 0.062 | 1.06 | 2.58E-09 | Bellenguez | Microglia | 0.008585298 |
| rs6742 | SLC2A4RG | 20 | 62374441 | C | T | 0.779 | 0.062 | 1.06 | 2.58E-09 | Bellenguez | Oligodendrocytes | 0.141434608 |
| rs6742 | SLC2A4RG | 20 | 62374441 | C | T | 0.779 | 0.062 | 1.06 | 2.58E-09 | Bellenguez | OPC | 0.0827254 |
| rs2154481 | APP | 21 | 27473875 | T | C | 0.524 | 0.041 | 1.04 | 1E-12 | Bellenguez | Astrocytes | 0.014809285 |
| rs2154481 | APP | 21 | 27473875 | T | C | 0.524 | 0.041 | 1.04 | 1E-12 | Bellenguez | Endothelial | 0.05997471 |
| rs2154481 | APP | 21 | 27473875 | T | C | 0.524 | 0.041 | 1.04 | 1E-12 | Bellenguez | GABAergic | 0.217915636 |
| rs2154481 | APP | 21 | 27473875 | T | C | 0.524 | 0.041 | 1.04 | 1E-12 | Bellenguez | Glutamatergic | 0.15046021 |
| rs2154481 | APP | 21 | 27473875 | T | C | 0.524 | 0.041 | 1.04 | 1E-12 | Bellenguez | Microglia | 0.047875632 |
| rs2154481 | APP | 21 | 27473875 | T | C | 0.524 | 0.041 | 1.04 | 1E-12 | Bellenguez | Oligodendrocytes | 0.339328385 |
| rs2154481 | APP | 21 | 27473875 | T | C | 0.524 | 0.041 | 1.04 | 1E-12 | Bellenguez | OPC | 0.169636142 |
| rs2830489 | ADAMTS1 | 21 | 28148191 | C | T | 0.719 | 0.030 | 1.03 | 1.69E-10 | Bellenguez | Astrocytes | 0.03088711 |
| rs2830489 | ADAMTS1 | 21 | 28148191 | C | T | 0.719 | 0.030 | 1.03 | 1.69E-10 | Bellenguez | Endothelial | 0 |

| | | | | | | | | | | | | |
|---|---|---|---|---|---|---|---|---|---|---|---|---|
| rs2830489 | ADAMTS1 | 21 | 28148191 | C | T | 0.719 | 0.030 | 1.03 | 1.69E-10 | Bellenguez | GABAergic | 0.106944476 |
| rs2830489 | ADAMTS1 | 21 | 28148191 | C | T | 0.719 | 0.030 | 1.03 | 1.69E-10 | Bellenguez | Glutamatergic | 0.025872192 |
| rs2830489 | ADAMTS1 | 21 | 28148191 | C | T | 0.719 | 0.030 | 1.03 | 1.69E-10 | Bellenguez | Microglia | 0.000682325 |
| rs2830489 | ADAMTS1 | 21 | 28148191 | C | T | 0.719 | 0.030 | 1.03 | 1.69E-10 | Bellenguez | Oligodendrocytes | 0.711336629 |
| rs2830489 | ADAMTS1 | 21 | 28148191 | C | T | 0.719 | 0.030 | 1.03 | 1.69E-10 | Bellenguez | OPC | 0.124277269 |
| rs143080277 | NCK2 | 2 | 106366056 | C | T | 0.005 | 0.365 | 1.44 | 2.07E-13 | Bellenguez | Astrocytes | 0.088353031 |
| rs143080277 | NCK2 | 2 | 106366056 | C | T | 0.005 | 0.365 | 1.44 | 2.07E-13 | Bellenguez | Endothelial | 0 |
| rs143080277 | NCK2 | 2 | 106366056 | C | T | 0.005 | 0.365 | 1.44 | 2.07E-13 | Bellenguez | GABAergic | 0.071826811 |
| rs143080277 | NCK2 | 2 | 106366056 | C | T | 0.005 | 0.365 | 1.44 | 2.07E-13 | Bellenguez | Glutamatergic | 0.11424498 |
| rs143080277 | NCK2 | 2 | 106366056 | C | T | 0.005 | 0.365 | 1.44 | 2.07E-13 | Bellenguez | Microglia | 0.586922488 |
| rs143080277 | NCK2 | 2 | 106366056 | C | T | 0.005 | 0.365 | 1.44 | 2.07E-13 | Bellenguez | Oligodendrocytes | 0.051248668 |
| rs143080277 | NCK2 | 2 | 106366056 | C | T | 0.005 | 0.365 | 1.44 | 2.07E-13 | Bellenguez | OPC | 0.087404021 |
| rs6733839 | BIN1 | 2 | 127892810 | T | C | 0.389 | 0.148 | 1.16 | 6.06E-118 | Bellenguez | Astrocytes | 0.01673033 |
| rs6733839 | BIN1 | 2 | 127892810 | T | C | 0.389 | 0.148 | 1.16 | 6.06E-118 | Bellenguez | Endothelial | 0.02169685 |
| rs6733839 | BIN1 | 2 | 127892810 | T | C | 0.389 | 0.148 | 1.16 | 6.06E-118 | Bellenguez | GABAergic | 0.109634502 |
| rs6733839 | BIN1 | 2 | 127892810 | T | C | 0.389 | 0.148 | 1.16 | 6.06E-118 | Bellenguez | Glutamatergic | 0.096723352 |
| rs6733839 | BIN1 | 2 | 127892810 | T | C | 0.389 | 0.148 | 1.16 | 6.06E-118 | Bellenguez | Microglia | 0.328455736 |
| rs6733839 | BIN1 | 2 | 127892810 | T | C | 0.389 | 0.148 | 1.16 | 6.06E-118 | Bellenguez | Oligodendrocytes | 0.410012315 |
| rs6733839 | BIN1 | 2 | 127892810 | T | C | 0.389 | 0.148 | 1.16 | 6.06E-118 | Bellenguez | OPC | 0.016746915 |
| rs10933431 | INPP5D | 2 | 233981912 | C | G | 0.766 | 0.041 | 1.04 | 3.62E-18 | Bellenguez | Astrocytes | 0.014098226 |
| rs10933431 | INPP5D | 2 | 233981912 | C | G | 0.766 | 0.041 | 1.04 | 3.62E-18 | Bellenguez | Endothelial | 0.152563232 |
| rs10933431 | INPP5D | 2 | 233981912 | C | G | 0.766 | 0.041 | 1.04 | 3.62E-18 | Bellenguez | GABAergic | 0.000696781 |
| rs10933431 | INPP5D | 2 | 233981912 | C | G | 0.766 | 0.041 | 1.04 | 3.62E-18 | Bellenguez | Glutamatergic | 0.004660807 |
| rs10933431 | INPP5D | 2 | 233981912 | C | G | 0.766 | 0.041 | 1.04 | 3.62E-18 | Bellenguez | Microglia | 0.823573842 |
| rs10933431 | INPP5D | 2 | 233981912 | C | G | 0.766 | 0.041 | 1.04 | 3.62E-18 | Bellenguez | Oligodendrocytes | 0.004327169 |
| rs10933431 | INPP5D | 2 | 233981912 | C | G | 0.766 | 0.041 | 1.04 | 3.62E-18 | Bellenguez | OPC | 7.99426E-05 |
| rs7597763 | INPP5D | 2 | 234082577 | C | A | 0.45 | 0.009 | 1.01 | 4.65E-09 | Wightman | Astrocytes | 0.014098226 |
| rs7597763 | INPP5D | 2 | 234082577 | C | A | 0.45 | 0.009 | 1.01 | 4.65E-09 | Wightman | Endothelial | 0.152563232 |
| rs7597763 | INPP5D | 2 | 234082577 | C | A | 0.45 | 0.009 | 1.01 | 4.65E-09 | Wightman | GABAergic | 0.000696781 |
| rs7597763 | INPP5D | 2 | 234082577 | C | A | 0.45 | 0.009 | 1.01 | 4.65E-09 | Wightman | Glutamatergic | 0.004660807 |
| rs7597763 | INPP5D | 2 | 234082577 | C | A | 0.45 | 0.009 | 1.01 | 4.65E-09 | Wightman | Microglia | 0.823573842 |
| rs7597763 | INPP5D | 2 | 234082577 | C | A | 0.45 | 0.009 | 1.01 | 4.65E-09 | Wightman | Oligodendrocytes | 0.004327169 |
| rs7597763 | INPP5D | 2 | 234082577 | C | A | 0.45 | 0.009 | 1.01 | 4.65E-09 | Wightman | OPC | 7.99426E-05 |
| rs17020490 | PRKD3 | 2 | 37531939 | C | T | 0.145 | 0.058 | 1.06 | 3.29E-09 | Bellenguez | Astrocytes | 0.069640688 |
| rs17020490 | PRKD3 | 2 | 37531939 | C | T | 0.145 | 0.058 | 1.06 | 3.29E-09 | Bellenguez | Endothelial | 0.339382413 |

| | | | | | | | | | | | | |
|---|---|---|---|---|---|---|---|---|---|---|---|---|
| rs17020490 | PRKD3 | 2 | 37531939 | C | T | 0.145 | 0.058 | 1.06 | 3.29E-09 | Bellenguez | GABAergic | 0.031766157 |
| rs17020490 | PRKD3 | 2 | 37531939 | C | T | 0.145 | 0.058 | 1.06 | 3.29E-09 | Bellenguez | Glutamatergic | 0.04274221 |
| rs17020490 | PRKD3 | 2 | 37531939 | C | T | 0.145 | 0.058 | 1.06 | 3.29E-09 | Bellenguez | Microglia | 0.392491005 |
| rs17020490 | PRKD3 | 2 | 37531939 | C | T | 0.145 | 0.058 | 1.06 | 3.29E-09 | Bellenguez | Oligodendrocytes | 0.0173526 |
| rs17020490 | PRKD3 | 2 | 37531939 | C | T | 0.145 | 0.058 | 1.06 | 3.29E-09 | Bellenguez | OPC | 0.106624927 |
| rs72777026 | ADAM17 | 2 | 9699011 | G | A | 0.144 | 0.058 | 1.06 | 2.72E-08 | Bellenguez | Astrocytes | 0.069554395 |
| rs72777026 | ADAM17 | 2 | 9699011 | G | A | 0.144 | 0.058 | 1.06 | 2.72E-08 | Bellenguez | Endothelial | 0.222712363 |
| rs72777026 | ADAM17 | 2 | 9699011 | G | A | 0.144 | 0.058 | 1.06 | 2.72E-08 | Bellenguez | GABAergic | 0.106182892 |
| rs72777026 | ADAM17 | 2 | 9699011 | G | A | 0.144 | 0.058 | 1.06 | 2.72E-08 | Bellenguez | Glutamatergic | 0.149197533 |
| rs72777026 | ADAM17 | 2 | 9699011 | G | A | 0.144 | 0.058 | 1.06 | 2.72E-08 | Bellenguez | Microglia | 0.144766662 |
| rs72777026 | ADAM17 | 2 | 9699011 | G | A | 0.144 | 0.058 | 1.06 | 2.72E-08 | Bellenguez | Oligodendrocytes | 0.151925267 |
| rs72777026 | ADAM17 | 2 | 9699011 | G | A | 0.144 | 0.058 | 1.06 | 2.72E-08 | Bellenguez | OPC | 0.155660886 |
| rs16824536 | MME | 3 | 154787511 | G | A | 0.946 | 0.083 | 1.09 | 3.63E-08 | Bellenguez | Astrocytes | 0.017877433 |
| rs16824536 | MME | 3 | 154787511 | G | A | 0.946 | 0.083 | 1.09 | 3.63E-08 | Bellenguez | Endothelial | 0 |
| rs16824536 | MME | 3 | 154787511 | G | A | 0.946 | 0.083 | 1.09 | 3.63E-08 | Bellenguez | GABAergic | 0.845255221 |
| rs16824536 | MME | 3 | 154787511 | G | A | 0.946 | 0.083 | 1.09 | 3.63E-08 | Bellenguez | Glutamatergic | 0.136867345 |
| rs16824536 | MME | 3 | 154787511 | G | A | 0.946 | 0.083 | 1.09 | 3.63E-08 | Bellenguez | Microglia | 0 |
| rs16824536 | MME | 3 | 154787511 | G | A | 0.946 | 0.083 | 1.09 | 3.63E-08 | Bellenguez | Oligodendrocytes | 0 |
| rs16824536 | MME | 3 | 154787511 | G | A | 0.946 | 0.083 | 1.09 | 3.63E-08 | Bellenguez | OPC | 0 |
| rs61762319 | MME | 3 | 154801978 | G | A | 0.026 | 0.166 | 1.18 | 2.16E-11 | Bellenguez | Astrocytes | 0.017877433 |
| rs61762319 | MME | 3 | 154801978 | G | A | 0.026 | 0.166 | 1.18 | 2.16E-11 | Bellenguez | Endothelial | 0 |
| rs61762319 | MME | 3 | 154801978 | G | A | 0.026 | 0.166 | 1.18 | 2.16E-11 | Bellenguez | GABAergic | 0.845255221 |
| rs61762319 | MME | 3 | 154801978 | G | A | 0.026 | 0.166 | 1.18 | 2.16E-11 | Bellenguez | Glutamatergic | 0.136867345 |
| rs61762319 | MME | 3 | 154801978 | G | A | 0.026 | 0.166 | 1.18 | 2.16E-11 | Bellenguez | Microglia | 0 |
| rs61762319 | MME | 3 | 154801978 | G | A | 0.026 | 0.166 | 1.18 | 2.16E-11 | Bellenguez | Oligodendrocytes | 0 |
| rs61762319 | MME | 3 | 154801978 | G | A | 0.026 | 0.166 | 1.18 | 2.16E-11 | Bellenguez | OPC | 0 |
| rs6846529 | HS3ST1 | 4 | 11025131 | C | T | 0.283 | 0.058 | 1.06 | 2.2E-17 | Bellenguez | Astrocytes | 6.44387E-05 |
| rs6846529 | HS3ST1 | 4 | 11025131 | C | T | 0.283 | 0.058 | 1.06 | 2.2E-17 | Bellenguez | Endothelial | 0.423686477 |
| rs6846529 | HS3ST1 | 4 | 11025131 | C | T | 0.283 | 0.058 | 1.06 | 2.2E-17 | Bellenguez | GABAergic | 0.131261239 |
| rs6846529 | HS3ST1 | 4 | 11025131 | C | T | 0.283 | 0.058 | 1.06 | 2.2E-17 | Bellenguez | Glutamatergic | 0.049953399 |
| rs6846529 | HS3ST1 | 4 | 11025131 | C | T | 0.283 | 0.058 | 1.06 | 2.2E-17 | Bellenguez | Microglia | 0.197060467 |
| rs6846529 | HS3ST1 | 4 | 11025131 | C | T | 0.283 | 0.058 | 1.06 | 2.2E-17 | Bellenguez | Oligodendrocytes | 0.013735253 |
| rs6846529 | HS3ST1 | 4 | 11025131 | C | T | 0.283 | 0.058 | 1.06 | 2.2E-17 | Bellenguez | OPC | 0.184238726 |
| rs2245466 | RHOH | 4 | 40198846 | G | C | 0.343 | 0.049 | 1.05 | 1.22E-09 | Bellenguez | Astrocytes | 0.00016661 |
| rs2245466 | RHOH | 4 | 40198846 | G | C | 0.343 | 0.049 | 1.05 | 1.22E-09 | Bellenguez | Endothelial | 0 |

| | | | | | | | | | | | | |
|---|---|---|---|---|---|---|---|---|---|---|---|---|
| rs2245466 | RHOH | 4 | 40198846 | G | C | 0.343 | 0.049 | 1.05 | 1.22E-09 | Bellenguez | GABAergic | 0.002458364 |
| rs2245466 | RHOH | 4 | 40198846 | G | C | 0.343 | 0.049 | 1.05 | 1.22E-09 | Bellenguez | Glutamatergic | 0.000591442 |
| rs2245466 | RHOH | 4 | 40198846 | G | C | 0.343 | 0.049 | 1.05 | 1.22E-09 | Bellenguez | Microglia | 0.974766195 |
| rs2245466 | RHOH | 4 | 40198846 | G | C | 0.343 | 0.049 | 1.05 | 1.22E-09 | Bellenguez | Oligodendrocytes | 0.000552742 |
| rs2245466 | RHOH | 4 | 40198846 | G | C | 0.343 | 0.049 | 1.05 | 1.22E-09 | Bellenguez | OPC | 0.021464648 |
| rs3822030 | IDUA | 4 | 987343 | T | G | 0.571 | 0.041 | 1.04 | 8.29E-12 | Bellenguez | Astrocytes | 0.090237488 |
| rs3822030 | IDUA | 4 | 987343 | T | G | 0.571 | 0.041 | 1.04 | 8.29E-12 | Bellenguez | Endothelial | 0.670594801 |
| rs3822030 | IDUA | 4 | 987343 | T | G | 0.571 | 0.041 | 1.04 | 8.29E-12 | Bellenguez | GABAergic | 0.037650915 |
| rs3822030 | IDUA | 4 | 987343 | T | G | 0.571 | 0.041 | 1.04 | 8.29E-12 | Bellenguez | Glutamatergic | 0.064575741 |
| rs3822030 | IDUA | 4 | 987343 | T | G | 0.571 | 0.041 | 1.04 | 8.29E-12 | Bellenguez | Microglia | 0.083270031 |
| rs3822030 | IDUA | 4 | 987343 | T | G | 0.571 | 0.041 | 1.04 | 8.29E-12 | Bellenguez | Oligodendrocytes | 0.022667417 |
| rs3822030 | IDUA | 4 | 987343 | T | G | 0.571 | 0.041 | 1.04 | 8.29E-12 | Bellenguez | OPC | 0.031003606 |
| rs112403360 | ANKH | 5 | 14724413 | A | T | 0.073 | 0.131 | 1.14 | 2.27E-09 | Bellenguez | Astrocytes | 0.040600581 |
| rs112403360 | ANKH | 5 | 14724413 | A | T | 0.073 | 0.131 | 1.14 | 2.27E-09 | Bellenguez | Endothelial | 0.171037782 |
| rs112403360 | ANKH | 5 | 14724413 | A | T | 0.073 | 0.131 | 1.14 | 2.27E-09 | Bellenguez | GABAergic | 0.256356472 |
| rs112403360 | ANKH | 5 | 14724413 | A | T | 0.073 | 0.131 | 1.14 | 2.27E-09 | Bellenguez | Glutamatergic | 0.126589538 |
| rs112403360 | ANKH | 5 | 14724413 | A | T | 0.073 | 0.131 | 1.14 | 2.27E-09 | Bellenguez | Microglia | 0.062234958 |
| rs112403360 | ANKH | 5 | 14724413 | A | T | 0.073 | 0.131 | 1.14 | 2.27E-09 | Bellenguez | Oligodendrocytes | 0.200819437 |
| rs112403360 | ANKH | 5 | 14724413 | A | T | 0.073 | 0.131 | 1.14 | 2.27E-09 | Bellenguez | OPC | 0.142361233 |
| rs871269 | TNIP1 | 5 | 150432388 | C | T | 0.674 | 0.041 | 1.04 | 8.67E-09 | Bellenguez | Astrocytes | 0.139933278 |
| rs871269 | TNIP1 | 5 | 150432388 | C | T | 0.674 | 0.041 | 1.04 | 8.67E-09 | Bellenguez | Endothelial | 0 |
| rs871269 | TNIP1 | 5 | 150432388 | C | T | 0.674 | 0.041 | 1.04 | 8.67E-09 | Bellenguez | GABAergic | 0.202411359 |
| rs871269 | TNIP1 | 5 | 150432388 | C | T | 0.674 | 0.041 | 1.04 | 8.67E-09 | Bellenguez | Glutamatergic | 0.174748626 |
| rs871269 | TNIP1 | 5 | 150432388 | C | T | 0.674 | 0.041 | 1.04 | 8.67E-09 | Bellenguez | Microglia | 0.180060733 |
| rs871269 | TNIP1 | 5 | 150432388 | C | T | 0.674 | 0.041 | 1.04 | 8.67E-09 | Bellenguez | Oligodendrocytes | 0.16484109 |
| rs871269 | TNIP1 | 5 | 150432388 | C | T | 0.674 | 0.041 | 1.04 | 8.67E-09 | Bellenguez | OPC | 0.138004915 |
| rs6891966 | HAVCR2 | 5 | 156526331 | G | A | 0.77 | 0.010 | 1.01 | 7.91E-10 | Wightman | Astrocytes | 0.004446692 |
| rs6891966 | HAVCR2 | 5 | 156526331 | G | A | 0.77 | 0.010 | 1.01 | 7.91E-10 | Wightman | Endothelial | 0.00035436 |
| rs6891966 | HAVCR2 | 5 | 156526331 | G | A | 0.77 | 0.010 | 1.01 | 7.91E-10 | Wightman | GABAergic | 0.005601434 |
| rs6891966 | HAVCR2 | 5 | 156526331 | G | A | 0.77 | 0.010 | 1.01 | 7.91E-10 | Wightman | Glutamatergic | 0.00790093 |
| rs6891966 | HAVCR2 | 5 | 156526331 | G | A | 0.77 | 0.010 | 1.01 | 7.91E-10 | Wightman | Microglia | 0.967503609 |
| rs6891966 | HAVCR2 | 5 | 156526331 | G | A | 0.77 | 0.010 | 1.01 | 7.91E-10 | Wightman | Oligodendrocytes | 0.012828822 |
| rs6891966 | HAVCR2 | 5 | 156526331 | G | A | 0.77 | 0.010 | 1.01 | 7.91E-10 | Wightman | OPC | 0.001364153 |
| rs113706587 | RASGEF1C | 5 | 179628150 | A | G | 0.11 | 0.086 | 1.09 | 2.22E-16 | Bellenguez | Astrocytes | 0.004818113 |
| rs113706587 | RASGEF1C | 5 | 179628150 | A | G | 0.11 | 0.086 | 1.09 | 2.22E-16 | Bellenguez | Endothelial | 0 |

| | | | | | | | | | | | | |
|---|---|---|---|---|---|---|---|---|---|---|---|---|
| rs113706587 | RASGEF1C | 5 | 179628150 | A | G | 0.11 | 0.086 | 1.09 | 2.22E-16 | Bellenguez | GABAergic | 0.017676801 |
| rs113706587 | RASGEF1C | 5 | 179628150 | A | G | 0.11 | 0.086 | 1.09 | 2.22E-16 | Bellenguez | Glutamatergic | 0.027452112 |
| rs113706587 | RASGEF1C | 5 | 179628150 | A | G | 0.11 | 0.086 | 1.09 | 2.22E-16 | Bellenguez | Microglia | 0.920178605 |
| rs113706587 | RASGEF1C | 5 | 179628150 | A | G | 0.11 | 0.086 | 1.09 | 2.22E-16 | Bellenguez | Oligodendrocytes | 0.015525203 |
| rs113706587 | RASGEF1C | 5 | 179628150 | A | G | 0.11 | 0.086 | 1.09 | 2.22E-16 | Bellenguez | OPC | 0.014349167 |
| rs62374257 | COX7C | 5 | 86223195 | C | T | 0.23 | 0.058 | 1.06 | 1.38E-15 | Bellenguez | Astrocytes | 0.267403697 |
| rs62374257 | COX7C | 5 | 86223195 | C | T | 0.23 | 0.058 | 1.06 | 1.38E-15 | Bellenguez | Endothelial | 0.004015617 |
| rs62374257 | COX7C | 5 | 86223195 | C | T | 0.23 | 0.058 | 1.06 | 1.38E-15 | Bellenguez | GABAergic | 0.277366517 |
| rs62374257 | COX7C | 5 | 86223195 | C | T | 0.23 | 0.058 | 1.06 | 1.38E-15 | Bellenguez | Glutamatergic | 0.19801639 |
| rs62374257 | COX7C | 5 | 86223195 | C | T | 0.23 | 0.058 | 1.06 | 1.38E-15 | Bellenguez | Microglia | 0.068002331 |
| rs62374257 | COX7C | 5 | 86223195 | C | T | 0.23 | 0.058 | 1.06 | 1.38E-15 | Bellenguez | Oligodendrocytes | 0.096795108 |
| rs62374257 | COX7C | 5 | 86223195 | C | T | 0.23 | 0.058 | 1.06 | 1.38E-15 | Bellenguez | OPC | 0.08840034 |
| rs785129 | HS3ST5 | 6 | 114612895 | T | C | 0.35 | 0.039 | 1.04 | 2.4E-09 | Bellenguez | Astrocytes | 0.064067733 |
| rs785129 | HS3ST5 | 6 | 114612895 | T | C | 0.35 | 0.039 | 1.04 | 2.4E-09 | Bellenguez | Endothelial | 0 |
| rs785129 | HS3ST5 | 6 | 114612895 | T | C | 0.35 | 0.039 | 1.04 | 2.4E-09 | Bellenguez | GABAergic | 0.253173003 |
| rs785129 | HS3ST5 | 6 | 114612895 | T | C | 0.35 | 0.039 | 1.04 | 2.4E-09 | Bellenguez | Glutamatergic | 0.069791917 |
| rs785129 | HS3ST5 | 6 | 114612895 | T | C | 0.35 | 0.039 | 1.04 | 2.4E-09 | Bellenguez | Microglia | 0.0001625 |
| rs785129 | HS3ST5 | 6 | 114612895 | T | C | 0.35 | 0.039 | 1.04 | 2.4E-09 | Bellenguez | Oligodendrocytes | 0.577803157 |
| rs785129 | HS3ST5 | 6 | 114612895 | T | C | 0.35 | 0.039 | 1.04 | 2.4E-09 | Bellenguez | OPC | 0.035001689 |
| rs6605556 | HLA-DQA1 | 6 | 32583099 | A | G | 0.839 | 0.062 | 1.06 | 7.07E-20 | Bellenguez | Astrocytes | 0.000300908 |
| rs6605556 | HLA-DQA1 | 6 | 32583099 | A | G | 0.839 | 0.062 | 1.06 | 7.07E-20 | Bellenguez | Endothelial | 0 |
| rs6605556 | HLA-DQA1 | 6 | 32583099 | A | G | 0.839 | 0.062 | 1.06 | 7.07E-20 | Bellenguez | GABAergic | 0.000763701 |
| rs6605556 | HLA-DQA1 | 6 | 32583099 | A | G | 0.839 | 0.062 | 1.06 | 7.07E-20 | Bellenguez | Glutamatergic | 0.002989408 |
| rs6605556 | HLA-DQA1 | 6 | 32583099 | A | G | 0.839 | 0.062 | 1.06 | 7.07E-20 | Bellenguez | Microglia | 0.99117136 |
| rs6605556 | HLA-DQA1 | 6 | 32583099 | A | G | 0.839 | 0.062 | 1.06 | 7.07E-20 | Bellenguez | Oligodendrocytes | 0.004774622 |
| rs6605556 | HLA-DQA1 | 6 | 32583099 | A | G | 0.839 | 0.062 | 1.06 | 7.07E-20 | Bellenguez | OPC | 0 |
| rs10947943 | UNC5CL | 6 | 41004093 | G | A | 0.858 | 0.073 | 1.08 | 1.13E-09 | Bellenguez | Astrocytes | 0.442416806 |
| rs10947943 | UNC5CL | 6 | 41004093 | G | A | 0.858 | 0.073 | 1.08 | 1.13E-09 | Bellenguez | Endothelial | 0.05647879 |
| rs10947943 | UNC5CL | 6 | 41004093 | G | A | 0.858 | 0.073 | 1.08 | 1.13E-09 | Bellenguez | GABAergic | 0.033140507 |
| rs10947943 | UNC5CL | 6 | 41004093 | G | A | 0.858 | 0.073 | 1.08 | 1.13E-09 | Bellenguez | Glutamatergic | 0.064060776 |
| rs10947943 | UNC5CL | 6 | 41004093 | G | A | 0.858 | 0.073 | 1.08 | 1.13E-09 | Bellenguez | Microglia | 0 |
| rs10947943 | UNC5CL | 6 | 41004093 | G | A | 0.858 | 0.073 | 1.08 | 1.13E-09 | Bellenguez | Oligodendrocytes | 0.402724753 |
| rs10947943 | UNC5CL | 6 | 41004093 | G | A | 0.858 | 0.073 | 1.08 | 1.13E-09 | Bellenguez | OPC | 0.001178367 |
| rs143332484 | TREM2 | 6 | 41129207 | T | C | 0.013 | 0.365 | 1.44 | 2.78E-25 | Bellenguez | Astrocytes | 0.000549831 |
| rs143332484 | TREM2 | 6 | 41129207 | T | C | 0.013 | 0.365 | 1.44 | 2.78E-25 | Bellenguez | Endothelial | 0 |

| | | | | | | | | | | | | |
|---|---|---|---|---|---|---|---|---|---|---|---|---|
| rs143332484 | TREM2 | 6 | 41129207 | T | C | 0.013 | 0.365 | 1.44 | 2.78E-25 | Bellenguez | GABAergic | 0.000156616 |
| rs143332484 | TREM2 | 6 | 41129207 | T | C | 0.013 | 0.365 | 1.44 | 2.78E-25 | Bellenguez | Glutamatergic | 0.000116756 |
| rs143332484 | TREM2 | 6 | 41129207 | T | C | 0.013 | 0.365 | 1.44 | 2.78E-25 | Bellenguez | Microglia | 0.99910791 |
| rs143332484 | TREM2 | 6 | 41129207 | T | C | 0.013 | 0.365 | 1.44 | 2.78E-25 | Bellenguez | Oligodendrocytes | 2.89366E-05 |
| rs143332484 | TREM2 | 6 | 41129207 | T | C | 0.013 | 0.365 | 1.44 | 2.78E-25 | Bellenguez | OPC | 3.99506E-05 |
| rs75932628 | TREM2 | 6 | 41129252 | T | C | 0.003 | 0.833 | 2.30 | 2.53E-37 | Bellenguez | Astrocytes | 0.000549831 |
| rs75932628 | TREM2 | 6 | 41129252 | T | C | 0.003 | 0.833 | 2.30 | 2.53E-37 | Bellenguez | Endothelial | 0 |
| rs75932628 | TREM2 | 6 | 41129252 | T | C | 0.003 | 0.833 | 2.30 | 2.53E-37 | Bellenguez | GABAergic | 0.000156616 |
| rs75932628 | TREM2 | 6 | 41129252 | T | C | 0.003 | 0.833 | 2.30 | 2.53E-37 | Bellenguez | Glutamatergic | 0.000116756 |
| rs75932628 | TREM2 | 6 | 41129252 | T | C | 0.003 | 0.833 | 2.30 | 2.53E-37 | Bellenguez | Microglia | 0.99910791 |
| rs75932628 | TREM2 | 6 | 41129252 | T | C | 0.003 | 0.833 | 2.30 | 2.53E-37 | Bellenguez | Oligodendrocytes | 2.89366E-05 |
| rs75932628 | TREM2 | 6 | 41129252 | T | C | 0.003 | 0.833 | 2.30 | 2.53E-37 | Bellenguez | OPC | 3.99506E-05 |
| rs60755019 | TREML2 | 6 | 41149008 | G | A | 0.004 | 0.412 | 1.51 | 2.07E-08 | Bellenguez | Astrocytes | 0 |
| rs60755019 | TREML2 | 6 | 41149008 | G | A | 0.004 | 0.412 | 1.51 | 2.07E-08 | Bellenguez | Endothelial | 0 |
| rs60755019 | TREML2 | 6 | 41149008 | G | A | 0.004 | 0.412 | 1.51 | 2.07E-08 | Bellenguez | GABAergic | 0.589078345 |
| rs60755019 | TREML2 | 6 | 41149008 | G | A | 0.004 | 0.412 | 1.51 | 2.07E-08 | Bellenguez | Glutamatergic | 0.143345917 |
| rs60755019 | TREML2 | 6 | 41149008 | G | A | 0.004 | 0.412 | 1.51 | 2.07E-08 | Bellenguez | Microglia | 0.267575738 |
| rs60755019 | TREML2 | 6 | 41149008 | G | A | 0.004 | 0.412 | 1.51 | 2.07E-08 | Bellenguez | Oligodendrocytes | 0 |
| rs60755019 | TREML2 | 6 | 41149008 | G | A | 0.004 | 0.412 | 1.51 | 2.07E-08 | Bellenguez | OPC | 0 |
| rs7767350 | CD2AP | 6 | 47485126 | T | C | 0.271 | 0.095 | 1.10 | 7.94E-22 | Bellenguez | Astrocytes | 0.0620916 |
| rs7767350 | CD2AP | 6 | 47485126 | T | C | 0.271 | 0.095 | 1.10 | 7.94E-22 | Bellenguez | Endothelial | 0.329647831 |
| rs7767350 | CD2AP | 6 | 47485126 | T | C | 0.271 | 0.095 | 1.10 | 7.94E-22 | Bellenguez | GABAergic | 0.075734229 |
| rs7767350 | CD2AP | 6 | 47485126 | T | C | 0.271 | 0.095 | 1.10 | 7.94E-22 | Bellenguez | Glutamatergic | 0.086146858 |
| rs7767350 | CD2AP | 6 | 47485126 | T | C | 0.271 | 0.095 | 1.10 | 7.94E-22 | Bellenguez | Microglia | 0.314614405 |
| rs7767350 | CD2AP | 6 | 47485126 | T | C | 0.271 | 0.095 | 1.10 | 7.94E-22 | Bellenguez | Oligodendrocytes | 0.038748593 |
| rs7767350 | CD2AP | 6 | 47485126 | T | C | 0.271 | 0.095 | 1.10 | 7.94E-22 | Bellenguez | OPC | 0.093016484 |
| rs13237518 | TMEM106B | 7 | 12269593 | C | A | 0.588 | 0.051 | 1.05 | 4.88E-11 | Bellenguez | Astrocytes | 0.063316889 |
| rs13237518 | TMEM106B | 7 | 12269593 | C | A | 0.588 | 0.051 | 1.05 | 4.88E-11 | Bellenguez | Endothelial | 0.164969802 |
| rs13237518 | TMEM106B | 7 | 12269593 | C | A | 0.588 | 0.051 | 1.05 | 4.88E-11 | Bellenguez | GABAergic | 0.239787038 |
| rs13237518 | TMEM106B | 7 | 12269593 | C | A | 0.588 | 0.051 | 1.05 | 4.88E-11 | Bellenguez | Glutamatergic | 0.150991372 |
| rs13237518 | TMEM106B | 7 | 12269593 | C | A | 0.588 | 0.051 | 1.05 | 4.88E-11 | Bellenguez | Microglia | 0.040848263 |
| rs13237518 | TMEM106B | 7 | 12269593 | C | A | 0.588 | 0.051 | 1.05 | 4.88E-11 | Bellenguez | Oligodendrocytes | 0.181473196 |
| rs13237518 | TMEM106B | 7 | 12269593 | C | A | 0.588 | 0.051 | 1.05 | 4.88E-11 | Bellenguez | OPC | 0.15861344 |
| rs11771145 | EPHA1 | 7 | 143110762 | G | A | 0.652 | 0.041 | 1.04 | 3.3E-14 | Bellenguez | Astrocytes | 0.000495191 |
| rs11771145 | EPHA1 | 7 | 143110762 | G | A | 0.652 | 0.041 | 1.04 | 3.3E-14 | Bellenguez | Endothelial | 0.952165655 |

| | | | | | | | | | | | | |
|---|---|---|---|---|---|---|---|---|---|---|---|---|
| rs11771145 | EPHA1 | 7 | 143110762 | G | A | 0.652 | 0.041 | 1.04 | 3.3E-14 | Bellenguez | GABAergic | 0.005255062 |
| rs11771145 | EPHA1 | 7 | 143110762 | G | A | 0.652 | 0.041 | 1.04 | 3.3E-14 | Bellenguez | Glutamatergic | 0.040877569 |
| rs11771145 | EPHA1 | 7 | 143110762 | G | A | 0.652 | 0.041 | 1.04 | 3.3E-14 | Bellenguez | Microglia | 0 |
| rs11771145 | EPHA1 | 7 | 143110762 | G | A | 0.652 | 0.041 | 1.04 | 3.3E-14 | Bellenguez | Oligodendrocytes | 0.001206523 |
| rs11771145 | EPHA1 | 7 | 143110762 | G | A | 0.652 | 0.041 | 1.04 | 3.3E-14 | Bellenguez | OPC | 0 |
| rs6966331 | EPDR1 | 7 | 37883793 | C | T | 0.651 | 0.051 | 1.05 | 4.64E-10 | Bellenguez | Astrocytes | 0.094879582 |
| rs6966331 | EPDR1 | 7 | 37883793 | C | T | 0.651 | 0.051 | 1.05 | 4.64E-10 | Bellenguez | Endothelial | 0.147765302 |
| rs6966331 | EPDR1 | 7 | 37883793 | C | T | 0.651 | 0.051 | 1.05 | 4.64E-10 | Bellenguez | GABAergic | 0.310591319 |
| rs6966331 | EPDR1 | 7 | 37883793 | C | T | 0.651 | 0.051 | 1.05 | 4.64E-10 | Bellenguez | Glutamatergic | 0.24631584 |
| rs6966331 | EPDR1 | 7 | 37883793 | C | T | 0.651 | 0.051 | 1.05 | 4.64E-10 | Bellenguez | Microglia | 0.000269065 |
| rs6966331 | EPDR1 | 7 | 37883793 | C | T | 0.651 | 0.051 | 1.05 | 4.64E-10 | Bellenguez | Oligodendrocytes | 0.056371216 |
| rs6966331 | EPDR1 | 7 | 37883793 | C | T | 0.651 | 0.051 | 1.05 | 4.64E-10 | Bellenguez | OPC | 0.143807676 |
| rs76928645 | SEC61G | 7 | 54941328 | C | T | 0.897 | 0.062 | 1.06 | 1.62E-10 | Bellenguez | Astrocytes | 0.130176603 |
| rs76928645 | SEC61G | 7 | 54941328 | C | T | 0.897 | 0.062 | 1.06 | 1.62E-10 | Bellenguez | Endothelial | 0 |
| rs76928645 | SEC61G | 7 | 54941328 | C | T | 0.897 | 0.062 | 1.06 | 1.62E-10 | Bellenguez | GABAergic | 0.343973145 |
| rs76928645 | SEC61G | 7 | 54941328 | C | T | 0.897 | 0.062 | 1.06 | 1.62E-10 | Bellenguez | Glutamatergic | 0.169781788 |
| rs76928645 | SEC61G | 7 | 54941328 | C | T | 0.897 | 0.062 | 1.06 | 1.62E-10 | Bellenguez | Microglia | 0.055968075 |
| rs76928645 | SEC61G | 7 | 54941328 | C | T | 0.897 | 0.062 | 1.06 | 1.62E-10 | Bellenguez | Oligodendrocytes | 0.149572412 |
| rs76928645 | SEC61G | 7 | 54941328 | C | T | 0.897 | 0.062 | 1.06 | 1.62E-10 | Bellenguez | OPC | 0.150527976 |
| rs6943429 | UMAD1 | 7 | 7856894 | T | C | 0.421 | 0.049 | 1.05 | 1.03E-10 | Bellenguez | Astrocytes | 0.039639714 |
| rs6943429 | UMAD1 | 7 | 7856894 | T | C | 0.421 | 0.049 | 1.05 | 1.03E-10 | Bellenguez | Endothelial | 0.202630281 |
| rs6943429 | UMAD1 | 7 | 7856894 | T | C | 0.421 | 0.049 | 1.05 | 1.03E-10 | Bellenguez | GABAergic | 0.168432004 |
| rs6943429 | UMAD1 | 7 | 7856894 | T | C | 0.421 | 0.049 | 1.05 | 1.03E-10 | Bellenguez | Glutamatergic | 0.101482756 |
| rs6943429 | UMAD1 | 7 | 7856894 | T | C | 0.421 | 0.049 | 1.05 | 1.03E-10 | Bellenguez | Microglia | 0.096806195 |
| rs6943429 | UMAD1 | 7 | 7856894 | T | C | 0.421 | 0.049 | 1.05 | 1.03E-10 | Bellenguez | Oligodendrocytes | 0.231385836 |
| rs6943429 | UMAD1 | 7 | 7856894 | T | C | 0.421 | 0.049 | 1.05 | 1.03E-10 | Bellenguez | OPC | 0.159623213 |
| rs10952097 | ICA1 | 7 | 8244012 | T | C | 0.114 | 0.068 | 1.07 | 6.81E-09 | Bellenguez | Astrocytes | 0.000704052 |
| rs10952097 | ICA1 | 7 | 8244012 | T | C | 0.114 | 0.068 | 1.07 | 6.81E-09 | Bellenguez | Endothelial | 0.079147847 |
| rs10952097 | ICA1 | 7 | 8244012 | T | C | 0.114 | 0.068 | 1.07 | 6.81E-09 | Bellenguez | GABAergic | 0.263115761 |
| rs10952097 | ICA1 | 7 | 8244012 | T | C | 0.114 | 0.068 | 1.07 | 6.81E-09 | Bellenguez | Glutamatergic | 0.322007942 |
| rs10952097 | ICA1 | 7 | 8244012 | T | C | 0.114 | 0.068 | 1.07 | 6.81E-09 | Bellenguez | Microglia | 0.084778848 |
| rs10952097 | ICA1 | 7 | 8244012 | T | C | 0.114 | 0.068 | 1.07 | 6.81E-09 | Bellenguez | Oligodendrocytes | 0.002368043 |
| rs10952097 | ICA1 | 7 | 8244012 | T | C | 0.114 | 0.068 | 1.07 | 6.81E-09 | Bellenguez | OPC | 0.247877507 |
| rs7384878 | SPDYE3 | 7 | 99932049 | T | C | 0.69 | 0.083 | 1.09 | 1.06E-26 | Bellenguez | Astrocytes | 0.058277231 |
| rs7384878 | SPDYE3 | 7 | 99932049 | T | C | 0.69 | 0.083 | 1.09 | 1.06E-26 | Bellenguez | Endothelial | 0.553676849 |

| | | | | | | | | | | | | |
|---|---|---|---|---|---|---|---|---|---|---|---|---|
| rs7384878 | SPDYE3 | 7 | 99932049 | T | C | 0.69 | 0.083 | 1.09 | 1.06E-26 | Bellenguez | GABAergic | 0.088419894 |
| rs7384878 | SPDYE3 | 7 | 99932049 | T | C | 0.69 | 0.083 | 1.09 | 1.06E-26 | Bellenguez | Glutamatergic | 0.078623505 |
| rs7384878 | SPDYE3 | 7 | 99932049 | T | C | 0.69 | 0.083 | 1.09 | 1.06E-26 | Bellenguez | Microglia | 0.038610528 |
| rs7384878 | SPDYE3 | 7 | 99932049 | T | C | 0.69 | 0.083 | 1.09 | 1.06E-26 | Bellenguez | Oligodendrocytes | 0.074778293 |
| rs7384878 | SPDYE3 | 7 | 99932049 | T | C | 0.69 | 0.083 | 1.09 | 1.06E-26 | Bellenguez | OPC | 0.1076137 |
| rs1065712 | CTSB | 8 | 11702122 | C | G | 0.053 | 0.058 | 1.06 | 1.94E-09 | Bellenguez | Astrocytes | 0.02527167 |
| rs1065712 | CTSB | 8 | 11702122 | C | G | 0.053 | 0.058 | 1.06 | 1.94E-09 | Bellenguez | Endothelial | 0.097170327 |
| rs1065712 | CTSB | 8 | 11702122 | C | G | 0.053 | 0.058 | 1.06 | 1.94E-09 | Bellenguez | GABAergic | 0.112987968 |
| rs1065712 | CTSB | 8 | 11702122 | C | G | 0.053 | 0.058 | 1.06 | 1.94E-09 | Bellenguez | Glutamatergic | 0.078240096 |
| rs1065712 | CTSB | 8 | 11702122 | C | G | 0.053 | 0.058 | 1.06 | 1.94E-09 | Bellenguez | Microglia | 0.597881374 |
| rs1065712 | CTSB | 8 | 11702122 | C | G | 0.053 | 0.058 | 1.06 | 1.94E-09 | Bellenguez | Oligodendrocytes | 0.02488416 |
| rs1065712 | CTSB | 8 | 11702122 | C | G | 0.053 | 0.058 | 1.06 | 1.94E-09 | Bellenguez | OPC | 0.063564405 |
| rs61732533 | OPLAH | 8 | 145108151 | A | G | 0.05 | 0.018 | 1.02 | 3.14E-09 | Wightman | Astrocytes | 0.575077474 |
| rs61732533 | OPLAH | 8 | 145108151 | A | G | 0.05 | 0.018 | 1.02 | 3.14E-09 | Wightman | Endothelial | 0 |
| rs61732533 | OPLAH | 8 | 145108151 | A | G | 0.05 | 0.018 | 1.02 | 3.14E-09 | Wightman | GABAergic | 0.018954212 |
| rs61732533 | OPLAH | 8 | 145108151 | A | G | 0.05 | 0.018 | 1.02 | 3.14E-09 | Wightman | Glutamatergic | 0.008838661 |
| rs61732533 | OPLAH | 8 | 145108151 | A | G | 0.05 | 0.018 | 1.02 | 3.14E-09 | Wightman | Microglia | 0.270534085 |
| rs61732533 | OPLAH | 8 | 145108151 | A | G | 0.05 | 0.018 | 1.02 | 3.14E-09 | Wightman | Oligodendrocytes | 0.110918331 |
| rs61732533 | OPLAH | 8 | 145108151 | A | G | 0.05 | 0.018 | 1.02 | 3.14E-09 | Wightman | OPC | 0.015677237 |
| rs34173062 | SHARPIN | 8 | 145158607 | A | G | 0.081 | 0.131 | 1.14 | 1.72E-16 | Bellenguez | Astrocytes | 0.128890195 |
| rs34173062 | SHARPIN | 8 | 145158607 | A | G | 0.081 | 0.131 | 1.14 | 1.72E-16 | Bellenguez | Endothelial | 0 |
| rs34173062 | SHARPIN | 8 | 145158607 | A | G | 0.081 | 0.131 | 1.14 | 1.72E-16 | Bellenguez | GABAergic | 0.083025979 |
| rs34173062 | SHARPIN | 8 | 145158607 | A | G | 0.081 | 0.131 | 1.14 | 1.72E-16 | Bellenguez | Glutamatergic | 0.052731307 |
| rs34173062 | SHARPIN | 8 | 145158607 | A | G | 0.081 | 0.131 | 1.14 | 1.72E-16 | Bellenguez | Microglia | 0.501565591 |
| rs34173062 | SHARPIN | 8 | 145158607 | A | G | 0.081 | 0.131 | 1.14 | 1.72E-16 | Bellenguez | Oligodendrocytes | 0.193374772 |
| rs34173062 | SHARPIN | 8 | 145158607 | A | G | 0.081 | 0.131 | 1.14 | 1.72E-16 | Bellenguez | OPC | 0.040412156 |
| rs73223431 | PTK2B | 8 | 27219987 | T | C | 0.369 | 0.068 | 1.07 | 4.03E-22 | Bellenguez | Astrocytes | 0.071616957 |
| rs73223431 | PTK2B | 8 | 27219987 | T | C | 0.369 | 0.068 | 1.07 | 4.03E-22 | Bellenguez | Endothelial | 0.185034896 |
| rs73223431 | PTK2B | 8 | 27219987 | T | C | 0.369 | 0.068 | 1.07 | 4.03E-22 | Bellenguez | GABAergic | 0.071344816 |
| rs73223431 | PTK2B | 8 | 27219987 | T | C | 0.369 | 0.068 | 1.07 | 4.03E-22 | Bellenguez | Glutamatergic | 0.454143245 |
| rs73223431 | PTK2B | 8 | 27219987 | T | C | 0.369 | 0.068 | 1.07 | 4.03E-22 | Bellenguez | Microglia | 0.122636234 |
| rs73223431 | PTK2B | 8 | 27219987 | T | C | 0.369 | 0.068 | 1.07 | 4.03E-22 | Bellenguez | Oligodendrocytes | 0.081745946 |
| rs73223431 | PTK2B | 8 | 27219987 | T | C | 0.369 | 0.068 | 1.07 | 4.03E-22 | Bellenguez | OPC | 0.013477907 |
| rs11787077 | CLU | 8 | 27465312 | C | T | 0.608 | 0.083 | 1.09 | 1.7E-44 | Bellenguez | Astrocytes | 0.479539054 |
| rs11787077 | CLU | 8 | 27465312 | C | T | 0.608 | 0.083 | 1.09 | 1.7E-44 | Bellenguez | Endothelial | 0.021997015 |

| | | | | | | | | | | | | |
|---|---|---|---|---|---|---|---|---|---|---|---|---|
| rs11787077 | CLU | 8 | 27465312 | C | T | 0.608 | 0.083 | 1.09 | 1.7E-44 | Bellenguez | GABAergic | 0.166718304 |
| rs11787077 | CLU | 8 | 27465312 | C | T | 0.608 | 0.083 | 1.09 | 1.7E-44 | Bellenguez | Glutamatergic | 0.091964371 |
| rs11787077 | CLU | 8 | 27465312 | C | T | 0.608 | 0.083 | 1.09 | 1.7E-44 | Bellenguez | Microglia | 0.040377341 |
| rs11787077 | CLU | 8 | 27465312 | C | T | 0.608 | 0.083 | 1.09 | 1.7E-44 | Bellenguez | Oligodendrocytes | 0.081220006 |
| rs11787077 | CLU | 8 | 27465312 | C | T | 0.608 | 0.083 | 1.09 | 1.7E-44 | Bellenguez | OPC | 0.118183909 |
| rs1800978 | ABCA1 | 9 | 107665978 | G | C | 0.13 | 0.039 | 1.04 | 1.59E-09 | Bellenguez | Astrocytes | 0.3036271 |
| rs1800978 | ABCA1 | 9 | 107665978 | G | C | 0.13 | 0.039 | 1.04 | 1.59E-09 | Bellenguez | Endothelial | 0.017584952 |
| rs1800978 | ABCA1 | 9 | 107665978 | G | C | 0.13 | 0.039 | 1.04 | 1.59E-09 | Bellenguez | GABAergic | 0.060419695 |
| rs1800978 | ABCA1 | 9 | 107665978 | G | C | 0.13 | 0.039 | 1.04 | 1.59E-09 | Bellenguez | Glutamatergic | 0.020799176 |
| rs1800978 | ABCA1 | 9 | 107665978 | G | C | 0.13 | 0.039 | 1.04 | 1.59E-09 | Bellenguez | Microglia | 0.221065559 |
| rs1800978 | ABCA1 | 9 | 107665978 | G | C | 0.13 | 0.039 | 1.04 | 1.59E-09 | Bellenguez | Oligodendrocytes | 0.037779174 |
| rs1800978 | ABCA1 | 9 | 107665978 | G | C | 0.13 | 0.039 | 1.04 | 1.59E-09 | Bellenguez | OPC | 0.338724344 |

rsid = rs number; Gene = nearest protein-coding gene; chr = chromosome; pos_hg19 = position (hg19); A1 = effect allele; A2 = other allele; A1freq = A1 frequency; b = beta; OR = odds ratio; p = p-value; source = source GWAS; Celltype = 1 of 7 brain cell types; Expression = relative expression level of the nearest gene across 7 brain cell types
OPC = oligodendrocyte precursor cells
*OR based on the Stage II analysis for SNPs from Bellenguez et al.

**Supplementary Table 3:** Association between PGS and tau-PET, amyloid-PET and cognitive performance

| Biomarkers | Longitudinal | | | |
|---|---|---|---|---|
| | n | β | *p* | *Adj. p* |
| **Tau-PET Braak 1** | 231 | 0.081 | 0.215 | 0.215 |
| **Tau-PET Braak 3+4** | 231 | 0.2 | 0.002 | 0.005 |
| **Tau-PET Braak 5+6** | 231 | 0.146 | 0.031 | 0.038 |
| **Aβ-PET centiloid** | 879 | 0.069 | 0.031 | 0.038 |
| **ADNI-MEM** | 1832 | -0.101 | <0.001 | <0.001 |
| **ADAS13** | 1813 | 0.083 | <0.001 | <0.001 |

Adj. p = FDR-adjusted p-value; β = standardized beta; p = p-val ; n = sample size

**Supplementary table 4:** Sample characteristics, stratified by amyloid status

| | Tau-PET | |
|---|---|---|
| | **Amyloid-positive (n=119)** | **Amyloid-negative (n=90)** |
| **Age** | 75.97 [58 - 92] | 72.69 [59 - 90] |
| **Sex (M/F)** | 61M/ 58F | 43M/ 47F |
| **Diagnosis (HC/MCI/AD)** | 66HC/ 38MCI/ 15AD | 61HC/ 25MCI/ 4AD |
| **APOE ε4 (-/+)** | 47/ 72 | 68/ 22 |
| **Ethnicity (% white)** | 90.80% | 85.60% |
| **Follow-up time, y** | 1.73 [0.65 - 3.95] | 1.86 [0.75 - 3.79] |

ADAS = Alzheimer's Disease Assessment Scale; APOE = apolipoprotein E; MEM = episodic memory, n = sample size.

Unless otherwise indicated, the summary statistics are presented as mean [range]

**Supplementary table 5:** Asssociation between PGS and change rate in tau-PET SUVRs stratified by amyloid-PET status

| Biomarkers | Amyloid-Positive | | | Amyloid-Negative | | |
|---|---|---|---|---|---|---|
| | **β** | ***p*** | **CI** | **β** | ***p*** | **CI** |
| **Tau-PET Braak 1** | 0.071 | 0.428 | [-0.11, 0.256] | 0.163 | 0.141 | [-0.058, 0.37] |
| **Tau-PET Braak 3+4** | 0.328 | <0.001 | [0.15, 0.479] | 0 | 0.999 | [-0.209, 0.222] |
| **Tau-PET Braak 5+6** | 0.356 | <0.001 | [0.205, 0.55] | -0.11 | 0.271 | [-0.309, 0.105] |

β = standardized beta; p = p-value; CI = Confidence intervals

**Supplementary table 6:** Asssociation between cell-type specific PGS and the rate of change in tau-PET, amyloid-PET and cognition

| Biomarkers | GABAergic neurons | | | Glutamatergic neurons | | | Microglia | | | Oligodendrocyes | | | oligodendrocyte precursor cells | | | Astrocytes | | | endothelial cells | | |
|---|---|---|---|---|---|---|---|---|---|---|---|---|---|---|---|---|---|---|---|---|---|
| | **β** | ***p*** | ***Adj. p*** | **β** | ***p*** | ***Adj. p*** | **β** | ***p*** | ***Adj. p*** | **β** | ***p*** | ***Adj. p*** | **β** | ***p*** | ***Adj. p*** | **β** | ***p*** | ***Adj. p*** | **β** | ***p*** | ***Adj. p*** |
| **Tau-PET Braak 1** | -0.067 | 0.308 | 0.479 | -0.042 | 0.531 | 0.62 | 0.101 | 0.124 | 0.26 | 0.069 | 0.301 | 0.479 | 0.034 | 0.603 | 0.649 | -0.061 | 0.364 | 0.513 | 0.048 | 0.466 | 0.594 |
| **Tau-PET Braak 3+4** | 0.052 | 0.439 | 0.576 | 0.062 | 0.366 | 0.513 | 0.16 | 0.015 | 0.043 | 0.153 | 0.023 | 0.056 | 0.055 | 0.401 | 0.543 | 0.021 | 0.756 | 0.756 | 0.036 | 0.584 | 0.649 |
| **Tau-PET Braak 5+6** | 0.082 | 0.231 | 0.428 | 0.097 | 0.166 | 0.331 | 0.08 | 0.239 | 0.428 | 0.146 | 0.034 | 0.079 | 0.021 | 0.752 | 0.756 | 0.076 | 0.27 | 0.454 | -0.033 | 0.63 | 0.662 |
| **Aβ-PET centiloid** | 0.02 | 0.532 | 0.62 | 0.037 | 0.245 | 0.428 | 0.074 | 0.02 | 0.053 | 0.059 | 0.064 | 0.142 | -0.022 | 0.491 | 0.606 | 0.017 | 0.597 | 0.649 | 0.03 | 0.349 | 0.513 |
| **ADNI-MEM** | -0.073 | <0.001 | 0.001 | -0.069 | <0.001 | 0.002 | -0.096 | <0.001 | <0.001 | -0.06 | 0.001 | 0.006 | -0.049 | 0.009 | 0.027 | -0.052 | 0.006 | 0.022 | -0.068 | <0.001 | 0.002 |
| **ADAS13** | 0.05 | 0.004 | 0.015 | 0.056 | 0.001 | 0.006 | 0.077 | <0.001 | <0.001 | 0.046 | 0.007 | 0.024 | 0.056 | 0.001 | 0.006 | 0.042 | 0.015 | 0.043 | 0.067 | <0.001 | 0.001 |

Adj. p = FDR-adjusted p-value; β = standardized beta; p = p-value

**Supplementary table 7:** Risk enrichment based on microglia PGS: Estimated sample size required for detecting intervention effects on tau-PET changes in the full sample and in a subgroup of Aβ+ individuals at power = 0.8

| | | Required number of participants per arm to detect intervention effect of: | |
|---|---|---|---|
| | | **20%** | **40%** |
| All | All participants | 630 | 144 |
| | APOE e4+ | 446 (29%) | 96 (33%) |
| | 4th Microglial PGS quartile | 543 (14%) | 125 (13%) |
| | 4th Microglial PGS quartile + APOE e4+ | 275 (56%) | 60 (58%) |
| Aβ+ | All Aβ+ participants | 440 | 97 |
| | APOE e4+ | 374 (15%) | 80 (18%) |
| | 4th Microglial PGS quartile | 284 (35%) | 63 (35%) |
| | 4th Microglial PGS quartile + APOE e4+ | 150 (66%) | 32 (67%) |

Percentage in brackets indicates the size of the sample size reduction relative to the reference group of no stratification (all participants) within either whole group (All) or the Aβ+ group.
APOE ε4+ = APOE ε3/ε4 & APOE ε4/ε4 carrier; 4th PGS quartile = group in upper quartile of residualized PGS scores; Aβ+ = abnormally high amyloid-PET accumulation

**Supplementary Table 8:** Polygenic Risk Score Reporting Standards (Wand et al. Nature, 591 (2021))

| | **PRS-RS criteria** | | | **PRS-RS criteria as applied by Rubinski et al (submitted)** | **Page number in Rubinski et al.** |
|---|---|---|---|---|---|
| ***Manuscript Section*** | ***Notes*** | ***PRS-RS Item*** | | | 3 |
| **Introduction** | Items listed in this section should set up the motivation and hypotheses of the polygenic risk score and integrated risk models. | Study Type | | Test the predictive value of PGS for the rate of change in tau-PET in Alzheimer's disease | 4 |
| | | Risk Model Purpose & Predicted Outcome | | PGS was used primarily for the prediction of the rate of change in tau-PET | 3 & 4 |
| **Methods** | Methods section should include information regarding the study participants' characteristics, including the distribution of these variables, variable definitions, and all features of the study that are relevant for interpretation. Methods should also detail all statistical models used or considered in the publication, including all parameters and assumptions. | Participants | Study Design & Recruitment | SNPs were selected from previous studies.<br>Analyses were performed based on longitudinal data from ADNI. | |
| | | | Demographic and Clinical Characteristics | ADNI demographics are presented in Table 1. | Table 1 |
| | | | Ancestry | SNPs were obtained from European-ansestry individuals.<br>Analyses were perefromed from ADNI participants which are primarily self-reporting as white. All analyses were controlled for self reported ethnicity and the first ten population components. | Table 1, p. 12 |
| | | Outcome of interest | | Rates of change in tau-PET, amyloid-PET and cognitive changes. | 16 |
| | | Non-Genetic Variables | | age, sex, years of education, diagnosis, self-reported ethnicity | 16 |
| | | Genetic data | | lead SNPs reported in Bellenguez et al., 2020 and Wightman et al., 2021, APOE e4 genotype | 14& Supplementary table 1 |
| | | Polygenic Risk Score Construction & Estimation | | The PGS score was calculated in PLINK (Chang et al., 2015) as the sum over the weighted number of alleles per SNP, using the respective log(OR) as weights; cell-type specific PGS: Variants were included if their closest gene exceeded a relative expression level of 0.1 across 7 cell-types | 14 |
| | | Integrated Risk Model | Model Type | PGS was modeled as continues variable. Models were controlled for age, sex, education, diagnosis, ethnicity, APOE genotype, the first ten principal components to correct for population stratification and maximum follow-up time. | 16 |
| | | | Model Fitting | Linear regression analysis | 16 |
| | | Missing Data | | For each statistical model, analysis was performed for individuals with full data, there were no missing data points. | NA |
| | | Statistical Methods | | Described in detail in the method section | 16 & 17 |
| | | Other Analyses | | Estimation of sample size needed | 17 |

| | | | | | |
|---|---|---|---|---|---|
| **Results** | Results should be presented for all conducted analyses, not just the final model, to enable full interpretation. This includes participant information (demographics, clinical characteristics, ancestry) of the training and test data, as well as all statistical models with relevant parameters. | Participants | Demographic and Clinical Characteristics | Available in Table 1, also sex-dependent effect is tested | Table 1 |
| | | | Ancestry | Ethnicity | Table 1 |
| | | PRS Distribution | | Available in Supplementary Figure 1 | Supplementary figure 1 |
| | | Risk Model Predictive Ability | | Predicive ability is described in detail in the results section | 5 - 7, supplementary tables 5 & 6 |
| | | Risk Model Discrimination | | Was not evaluated | NA |
| | | Risk Model Calibration | | Was not evaluated | NA |
| | | Subgroup Analyses | | Subgroup analyses were performed in samples stratified by Amyloid status. Demographics of subgroups are available in Supplementary table 4. | 6 |
| **Discussion** | The discussion should focus on the interpretation and contextualization of the risk model. This includes, but is not limited to, the generalizability, any limitations due to study characteristics or statistical assumptions, and the intended use case for the model. | Risk Model interpretation | | Risk model predicts tau accumulation also when controlled for APOE genotype, the major risk allel of AD.<br>PGS outperforms APOE in sample selection for clinical trials. | 9 |
| | | Limitations | | Described in detail in the discussion | 12 |
| | | Generalizability | | Our finding may not generalize to indviduals of other ethnic background. | 12 |
| | | Risk model Intended Uses | | May be used for prediction of tau progression, and for risk stratification in clinical trials. | 9 |
| **Transparency and Reproducibility** | Information related to the transparency and reproducibility of the study may not be included in the traditional main text, but rather in the data availability or funding statements at the end of a manuscript. | Data Availability | | The data that used in this study were obtained from the Alzheimer's disease Neuroimaging Initiative (ADNI) and are available from the ADNI database (adni.loni.usc.edu) upon registration and compliance with the data usage agreement. A source file for all figures as can be found in the supplementary. | 17 |
| | | Funding | | See manuscript | 23 |
